# Variations in the results of nutritional epidemiology studies due to analytic flexibility: Application of specification curve analysis to red meat and all-cause mortality

**DOI:** 10.1101/2023.12.19.23300248

**Authors:** Yumin Wang, Tyler Pitre, Joshua D. Wallach, Russell J. de Souza, Tanvir Jassal, Dennis Bier, Chirag J. Patel, Dena Zeraatkar

**Author notes:** **Corresponding author:** Dena Zeraatkar, 1280 Main St. W, Hamilton, Ontario. **Disclaimers:** Dr Wallach reported receiving grant support from the FDA, Arnold Ventures, Johnson & Johnson through Yale University, and the National Institute on Alcohol Abuse and Alcoholism of the NIH under award 1K01AA028258; serving as a consultant for Hagens Berman Sobol Shapiro LLP and Dugan Law Firm APLC; and serving as a medRxiv affiliate. **Funding**: None. **Contributions:** DZ and CJP conceptualized the study. DZ, YW, JDW, RJdS, TP, CJP, and DZ provided input on the study design and methods. YW, TJ, and TP collected data. YW performed analyses, with input from DZ and CJP. YW and DZ wrote the first draft of the manuscript and all authors provided critical comments. The senior author (manuscripts guarantor) affirms that the manuscript is an honest, accurate, and transparent account of the study being reported and that no important aspects of the study have been omitted. **Ethics approval:** Not required. **Patient/public engagement:** It was not possible to involve patients or the public in the design, conduct, reporting, or dissemination plans of our research.

## Abstract

**Objective:** To present an application of specification curve analysis—a novel analytic method that involves defining and implementing all plausible and valid analytic approaches for addressing a research question—to nutritional epidemiology.

**Data source:** National Health and Nutrition Examination Survey (NHANES) 2007 to 2014 linked with National Death Index.

**Methods:** We reviewed all observational studies addressing the effect of red meat on all-cause mortality, sourced from a published systematic review, and documented variations in analytic methods (e.g., choice of model, covariates, etc.). We enumerated all defensible combinations of analytic choices to produce a comprehensive list of all the ways in which the data may reasonably be analyzed. We applied specification curve analysis to NHANES data to investigate the effect of unprocessed red meat on all-cause mortality, using all reasonable analytic specifications.

**Results:** Among 15 publications reporting on 24 cohorts included in the systematic review on red meat and all-cause mortality, we identified 70 unique analytic methods, each including different analytic models, covariates, and operationalizations of red meat (e.g., continuous vs. quantiles). We applied specification curve analysis to NHANES, including 10,661 participants. Our specification curve analysis included 1,208 unique analytic specifications. Of 1,208 specifications, 435 (36.0%) yielded a hazard ratio equal to or above 1 for the effect of red meat on all-cause mortality and 773 (64.0%) below 1, with a median hazard ratio of 0.94 [IQR: 0.83 to 1.05]. Forty-eight specifications (3.97%) were statistically significant, 40 of which indicated unprocessed red meat to reduce all-cause mortality and 8 of which indicated red meat to increase mortality.

**Conclusion:** We show that the application of specification curve analysis to nutritional epidemiology is feasible and presents an innovative solution to analytic flexibility.

**Limitations:** Alternative analytic specifications may address slightly different questions and investigators may disagree about justifiable analytic approaches. Further, specification curve analysis is time and resource-intensive and may not always be feasible.

## Background

Unlike randomized trials for which investigators typically register protocols and statistical analysis plans before the collection of any data, when investigators analyze data from observational studies, there are often hundreds of equally justifiable ways of analyzing the data, each of which may produce results that vary in direction, magnitude, and statistical significance (1–7). The variability of effect estimates due to alternative analytical approaches is called ‘vibration of effects’ (2). Empirical evidence shows that results from observational studies may be highly dependent on analytic choices (1–5).

While our empirical and theoretical understanding of the question being investigated should guide our analytic choices, our knowledge of complex biomedical and environmental systems is limited and even experienced investigators come to different conclusions about the ideal analytic approach (4, 6, 8–13).

While we anticipate that discrepancies in analytic models often result from differences in opinions regarding the optimal analytic approach among well-intentioned investigators, some investigators may test many alternative analytic specifications and, intentionally or unintentionally, selectively report results for the specification that yields the most statistically significant or interesting results or results that support their preconceived hypotheses. Evidence shows that investigators’ prior beliefs and expectations influence their results (5). In the presence of very strong opinions, investigators’ beliefs and expectations may shape the literature to the detriment of empirical evidence (5).

### Nutritional epidemiology

Nutrition is a field particularly amenable to analytic flexibility (14). Trials investigating the health effects of nutritional exposures are often not feasible and so the evidence is primarily comprised of nutritional epidemiology studies—observational studies that recruit large groups of people and look for patterns between diet and health (15, 16).

The analysis of nutritional epidemiology data is complex and there is often limited consensus among experts about the ideal approach (17, 18). Sources of analytic flexibility include the type of analytic model (e.g., Poisson regression, Cox proportional hazards model), choice of covariates (i.e., investigators studying the same question will consider different adjusting variables (19)), operationalization of the exposure variable and covariates in the model (e.g., transformations, categorizations of continuous variables, functional form), and methods to address missing data, among others (8). Investigators often present several sensitivity analyses to investigate the effects of these uncertain analytic decisions on the results, but the choice of sensitivity analyses is also subjective and investigators may be more inclined to report sensitivity analyses that affirm their primary findings.

A large body of evidence shows inconsistency in the results of nutritional studies, some of which may be explained by analytic flexibility (3, 8, 20, 21). Such inconsistencies have eroded trust in nutritional epidemiology and subjected the field to criticism (22, 23). Nevertheless, nutritional epidemiology studies continue to play a crucial role in shaping dietary recommendations and policies, making it imperative to draw credible inferences from these studies (14, 15, 24).

### Specification curve analysis

Specification curve analysis—sometimes called multiverse analysis—is a novel analytic method that involves defining and implementing all plausible and valid analytic approaches for addressing a research question (25) (Box 1).

Through this approach, investigators define all plausible and justifiable choices for all aspects of the analysis (e.g., choice of model, covariates, etc.), enumerate all justifiable combinations of these choices to produce a comprehensive list of all the ways in which the data may be reasonably analyzed (i.e., analytic specifications), implement all or a random sample of the ways in which the question may be analyzed, and draw inferences using the distribution of results from all plausible analyses.

Specification curve analysis offers advantages to conventional methods for data analysis. It allows investigators to draw more credible inferences that are not contingent on arbitrary analytic decisions and reduces the opportunity for investigators to conduct many analyses and selectively report results for analyses that yield the most interesting results, though it does not completely eliminate subjectivity in analytic decisions.

While specification curve analysis has been previously applied in psychology and economics, it has seldom been applied in nutritional and environmental epidemiology (5, 26).

##### Box 1: Specification curve analysis

When investigators analyze data from observational studies, they may make numerous potentially justifiable, but still subjective, analytic decisions on which the direction, magnitude, and statistical significance of results may be contingent. Specification curve analysis may mitigate this issue (27).

Specification curve analysis involves defining and implementing all plausible and justifiable analytic methods for investigating a research question. Investigators subsequently interpret the distribution of results across all plausible analyses, instead of focusing on the results of only one analysis.

The implementation of specification curve analysis involves:

1. Defining all plausible choices across all aspects of the analysis. This typically includes:

- Criteria for selecting eligible participants for inclusion in the analysis
- Type of analytic model (e.g., logistic, Poisson, or Cox proportion hazards models)
- Choice of covariates
- Operationalizations of the exposure variable and covariates (e.g., transformations, functional form)
2. Enumerating all justifiable combinations of these analytic choices to to produce a comprehensive list of all the ways in which the data may be reasonably analyzed. For example, three unique choices for five aspects of the analysis yield 243 unique analytic specifications (=3 ).
3. Implementing all or a random sample of all reasonable analytic specifications.
4. Ordering the effect estimates from all analyses based on their direction and magnitude and presenting results on a specification curve plot. A specification curve plot reports the results of all analyses at the top and analytic characteristics at the bottom. The specification curve plot visually communicates the distribution of results across all specifications and the aspects of the analysis that are most consequential in influencing the direction and magnitude of findings.

### Objectives

We apply specification curve analysis to investigate the effect of unprocessed red meat on all-cause mortality—a question that has yielded inconsistent results in the literature and produced conflicting dietary recommendations.

A critical limitation of specification curve analysis is the subjectivity involved in selecting justifiable analytic specifications. Investigators may disagree about justifiable analytic approaches or may present results of analyses that are only marginally justifiable. To mitigate this issue, our analytic specifications were informed by the most common analytic methods used in previous published studies addressing the effects of red meat on all-cause mortality.

### Methods

This study was exempt from institutional ethics review because it uses secondary de-identified data. We report our results according to STROBE reporting guidelines for observational studies (28).

### Analytic specifications

We used a published systematic review of observational studies that addressed the effect of red meat on all-cause mortality to identify justifiable analytic specifications for specification curve analysis (29). We focus only on observational studies because randomized trials typically involve the preparation of detailed protocols and statistical analysis plans that reduce the analytic decisions available to investigators. While our objective was to investigate the effects of unprocessed red meat, we did not anticipate that studies investigating the effects of mixed unprocessed and processed red meat or unspecified types of red meat would use different analytic methods. Hence, we also reviewed studies that reported on mixed unprocessed and processed red meat and unspecified types of red meat.

Two reviewers, working independently and in duplicate, reviewed the primary studies from the systematic review and collected data on study characteristics and analytic methods, including the type of analytic model (e.g., Cox proportional hazards model, logistic regression), method of adjustment for energy (e.g., standard model, nutrient density model), covariates included in the model, operationalization of covariates (e.g., categorical, linear, quadratic), subgroup analyses (e.g., men vs. women), and the results of analyses, including secondary and sensitivity analyses, when reported. To ensure that the primary studies that we used to inform our analytic specifications addressed similar causal questions and interpreted their findings similarly, we documented the objectives of the primary studies and the ways in which the authors interpreted their findings.

### Study population

The National Health and Nutrition Examination Survey (NHANES) is a repeated cross-sectional probability survey by the US Centers for Disease Control and Prevention to characterize the health and nutritional status of the non-institutionalized, civilian US population (30). The survey is based on household interviews and physical examinations and is representative of the US population by its survey sampling method. The survey collects demographic, socioeconomic, dietary, and health-related data by household interview, and medical, dental, physiological measurements, and laboratory tests by physical examination.

For this analysis, we used the continuous 2007-2014 NHANES data linked with the National Death Index (31) and the Food Patterns Equivalents Database. The National Death Index is a database established by the National Center for Health Statistics (NCHS) that contains information on all deaths in the US. We extracted mortality status from the National Death Index up to 31 December 2019. The Food Patterns Equivalents Database contains information on the composition and nutritional content of individual foods.

We acknowledge that NHANES data is likely suboptimal compared to other nutrition datasets for investigating the effect of red meat and other nutritional exposures on health outcomes, due to it including few deaths and only collecting data on diet at a single point in time (30, 32). Our objective, however, is not to provide conclusive answers about the health effects of red meat but to demonstrate a proof-of-concept application of specification curve analysis to nutritional epidemiology. We used NHANES data due its availability to our team and our team’s familiarity with its structure.

We excluded participants with missing demographic, dietary, or lifestyle information; personal and family history of disease; pregnant people; and participants with implausible BMI (<15 or ≥60 kg/m ) or energy intake (<500kcal/day or >4,500kcal/day). To minimize missing data, we consolidated related variables in the database (e.g., when data was missing for the smoking history variable, we classified participants who endorsed smoking 0 cigarettes in their life as non-smokers).

Participants in NHANES completed two 24-hour dietary recalls, each conducted by trained interviewers and separated by 3-10 days, for which they provided information on intake of foods and beverages on each recall day (32). For our analysis, we define unprocessed red meat as any mammalian meat (i.e., beef, veal, pork, lamb, and game meat) (33).

### Data analysis

We performed specification curve analysis to investigate the effects of unprocessed red meat on all-cause mortality, using a Cox proportional hazards regression model with time since 24-hour recalls as the time variable in the model.

For each aspect of the analysis, we used the most used analytic choices from previous studies (Box 2) and enumerated all combinations of these choices to produce a comprehensive list of all plausible and reasonable analytic methods. We reviewed analytic specifications to confirm that every combination of analytic choices throughout the analysis was indeed justifiable. Although we intended to exclude specifications comprised of combinations that were not defensible, we found no such cases.

##### Box 2: Aspects of the analysis that varied across analytic specifications

1. Type of nutrition model

- Standard model
- Multivariable nutrient density model
2. Operationalization of red meat

- Continuous (per 100 g/day)
- Quartiles
- Quintiles
3. Subgroups of interest

- All participants
- Subgroup based on sex

- All females
- All males
- Subgroups based on age

- Participants aged 20-39 years old
- Participants aged 40-59 years old
- Participants 60-79 years old
4. Covariates

Aspects of the analysis that varied across primary studies included the type of nutrition model (i.e., standard model and multivariable nutrient density model), operationalization of red meat (i.e., continuous, quartiles, quintiles), subgroups of interest (i.e., only males, only females, 20-39 years old, 40-59 years old, 60-79 years old), and choice of covariates. The standard nutrition model adjusts for total energy in the analytic model while the multivariable nutrient density model includes total energy as a covariate and divides food intake by total energy intake (34). We did not consider the residual energy model since it is largely equivalent to the standard model (34).

We constructed two sets of covariates: covariates that we included in all models and covariates that were adjusted in some models. In all models, we adjusted for a core set of covariates that were considered in nearly all primary studies: age, sex, smoking, total energy intake, year, menopausal status, hormone therapy, parity, and oral contraceptives. We also optionally adjusted for a secondary set of other covariates that were only adjusted in some (but not all) studies: race/ethnicity (Mexican American/other Hispanic/non-Hispanic white/non-Hispanic black/other race–including multi-racial), education (less than 9^th^ grade/9-11 grade/high school graduate/some college or AA degree/college graduate or above), marital status, alcohol consumption, physical activity, BMI, socioeconomic status, comorbidities, and dietary variables.

We are unable to test for all possible combinations of covariates due to computational feasibility. Hence, we generated 20 unique combinations of covariates that all adjusted for the core set of variables and each of which adjusted for a random set of the secondary covariates. We applied specification curve analysis and computed HRs and 95% confidence intervals corresponding to the effect of red meat intake on all-cause mortality for each analytic specification.

For specifications in which red meat was treated as a continuous variable, we calculated HRs and associated confidence intervals corresponding to a 100 g/day increase in intake of red meat. For specifications in which red meat was treated as a categorical variable (e.g., quartiles or quintiles), we calculated hazard ratios and associated confidence intervals corresponding to the highest versus lowest quantile of red meat exposure. While these contrasts represent different quantities of red meat intake, primary observational nutritional epidemiology studies overlook these differences when interpreting results and systematic reviews and meta-analyses often combine these estimates from studies using disparate quantities (35). In our supplement, we present results stratified by how red meat is defined in analytic models (i.e., quartiles, quintiles, or continuous 100 g/day).

To test whether models from the specification curve analysis met the proportional hazards assumption, we selected a sample of all specifications at random and tested the correlation between Schoenfeld residuals and ranked failure time.

We excluded results from models that yielded what we considered to be implausible effect estimates (i.e., studies that yielded implausibly wide confidence intervals with lower bound HR ≤0.2 or upper bound HR ≥5). A review of analytic specifications that yielded results outside of this range suggested sparse data bias, where there are too few events in certain combinations of explanatory variables resulting in over- or under-estimation of effect estimates (36). While these thresholds are arbitrary, they pragmatically excluded specifications that yielded what we considered to be results beyond the range of effects we would plausibly expect from diet and nutrition on health outcomes.

We performed three statistical tests to address (i) whether the median effect estimate across all specifications is more extreme than would be expected if red meat had no effect on all-cause mortality, (ii) the proportion of specifications that produced statistically significant effects is more extreme than would expected if red meat had no effect on all-cause mortality, and (iii) whether Stouffer’s averaged Z value across all specifications is more extreme than would be expected if red meat had no effect on all-cause mortality (27). To perform these tests, we permuted red meat intake and sampled with replacement across all participants to yield 500 bootstrapped samples to which we applied specification curve analysis. Based on the results of the specification curve analysis to the permuted datasets, we calculate P-values using the percentage of bootstrap sample with results as or more extreme than the observed results. We used an alpha of 0.05 to indicate statistical significance.

We performed all analyses in R (Vienna, Austria; version 4.1.2), using the *specr* package for specification curve analysis (37). Data from NHANES is publicly accessible and the code to produce the results in this paper is available on a public repository: https://github.com/Yumin-Wang/Red-Meat-Consumption---All-Cause-Mortality.

## Results

### Study characteristics

A systematic review addressing the health effects of red meat identified 15 publications reporting on 24 cohort studies that examined the effect of red meat on all-cause mortality (29) (Supplement Table 1).

To ensure that these primary studies addressed similar causal questions and interpreted their findings similarly, we documented the objectives of the primary studies and the ways in which the authors interpreted their findings (Supplement Table 2). The primary aim of all except two of these studies was to investigate the effects of red meat on all-cause mortality. One study investigated the effects of substituting total and different types of dietary protein for carbohydrates on mortality but also presented models investigating the effects of isocaloric substitutions of carbohydrates for red meat on mortality (38). The second study investigated the effects of components of a traditional Sami diet, including red meat, on mortality (39).

Studies reported 70 unique methods to investigate the relationship between red meat and all-cause mortality (Supplement Tables 2 and 3). Studies varied in their choice of analytic model (e.g., Cox proportional hazards model, Poisson regression), adjustment for energy (e.g., standard model and nutrient density model), covariates included in the model, operationalizations of variables (e.g., functional form in the model), and subgroups. Typical studies performed time-dependent Cox regression models in which red meat was treated as a categorical variable in quartiles or quintiles and adjusted for age, sex, smoking, alcohol intake, physical activity, and BMI.

Studies reported relative effect estimates of red meat on all-cause mortality ranging between 0.63 to 2.31 (median: 1.14; IQR: 1.02 to 1.23). Supplement Figure 1 presents the results of the analyses reported in studies.

### Participant characteristics

We used data from NHANES 2007 to 2014 and excluded participants without mortality data and missing or implausible data, leaving 10,661 eligible participants.

Table 1 and Supplement Table 5 present participant characteristics. Our study included participants ranging from young adults to the elderly, with approximately equal representation of men and women. Most participants were white, non- or light smokers, with a median intake of unprocessed red meat less than half a serving per day. Figure 1 presents the selection of participants in the analysis.

**Figure 1:**
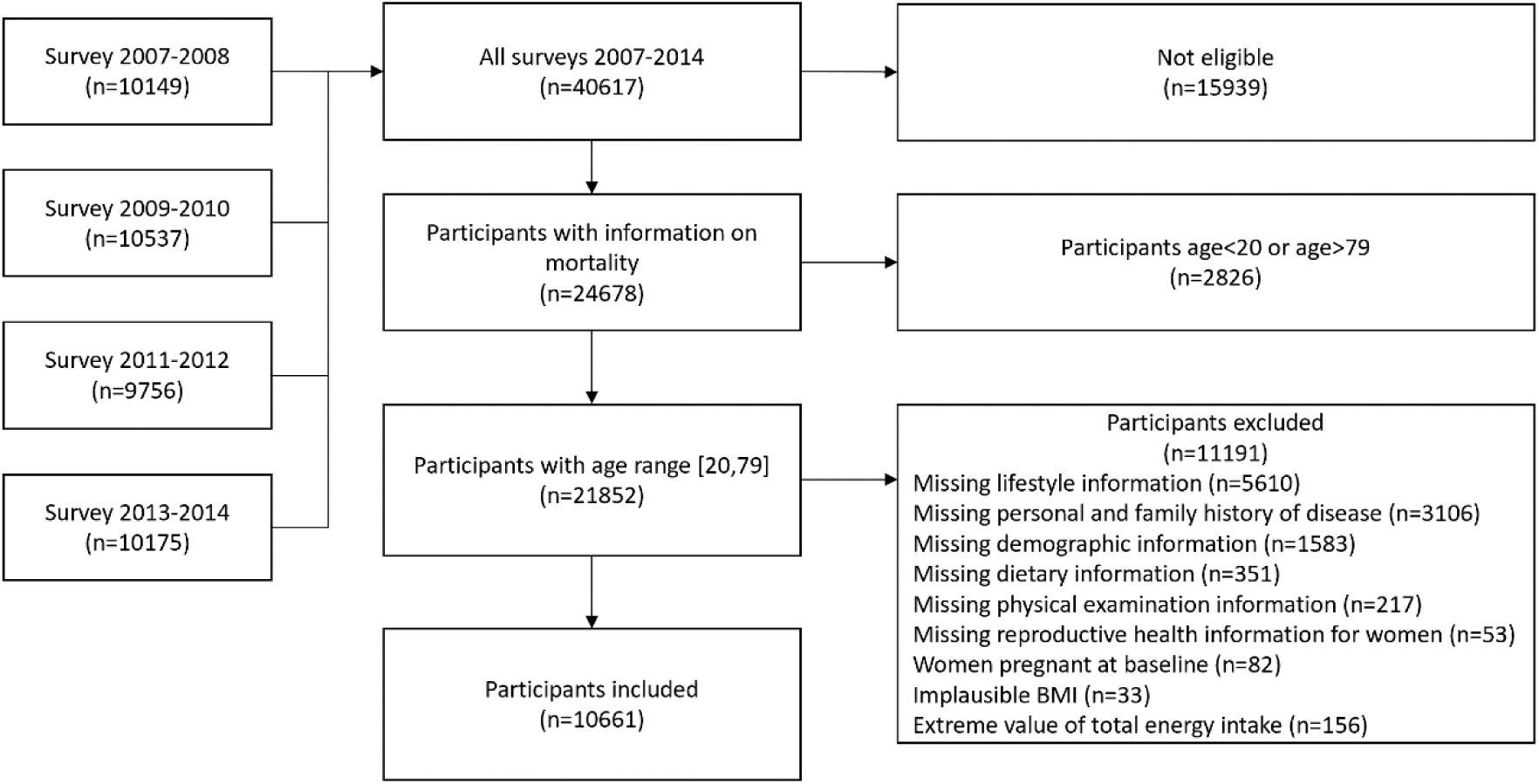
Selection of study participants from the National Health and Nutrition Examination Survey (NHANES) for inclusion in the analysis

**Table 1:** Participant characteristics.

|  |  |
| --- | --- |
| **Total participants, N** | 10,661 |
| **All-Cause mortality, n (%)** | 1022 (10) |
| **Follow-up (months)** | 99 (65, 143) |
| **Age (years)** | 50 (27, 71) |
| **Gender** |  |
| Female, n (%) | 5150 (48) |
| Male, n (%) | 5511 (52) |
| **Dietary Intakes** |  |
| Unprocessed red meat (g/d) | 29.5 (0, 120.2) |
| Total energy intake (kcal/d) | 1945 (1168, 3099) |
| **Years of entering cohort** |  |
| 2007-2008, n (%) | 2311 (22) |
| 2009-2010, n (%) | 2358 (22) |
| 2011-2012, n (%) | 2857 (27) |
| 2013-2014, n (%) | 3135 (29) |
| **Race/Ethnicity** |  |
| Mexican American, n (%) | 1321 (12) |
| Other Hispanic, n (%) | 988 (9) |
| Non-Hispanic White, n (%) | 5193 (49) |
| Non-Hispanic Black, n (%) | 2235 (21) |
| Other Race – Including Multi-Racial, n (%) | 924 (9) |
| **Smoking** |  |
| Non or light smoker, n (%) | 8373 (79) |
| Moderate smoker, n (%) | 437 (4) |
| Heavy smoker, n (%) | 1851 (17) |
| **BMI (kg/m<sup>2</sup>)** | 28.4 (21.9, 38.5) |

*Data presented as numbers and proportions or as medians (10<sup>th</sup> percentile, 90<sup>th</sup> percentile).*

### Specification curve analysis

Using all analytic choices identified in the primary studies, we enumerated all justifiable ways in which the data may be reasonably analyzed, yielding 1,440 unique analytic specifications. We were able to accommodate most analytic choices reported in primary studies using data from NHANES (Supplement Tables 2 and 3). We were unable to implement time-varying variables due to the cross-sectional nature of the NHANES data.

We implemented all 1,440 reasonable specifications and identified 1,208 unique specifications with plausible results and 232 with implausibly wide confidence intervals (lower bound HR ≤0.2 or upper bound HR ≥5). These implausible specifications occurred in analyses of subgroups of the total study population that included many adjusting covariates, suggesting sparse data bias (36).

Figure 2 presents the results of the specification curve analysis. Our specification curve analysis produced a median hazard ratio of 0.94 [IQR: 0.83 to 1.05] for the effect of red meat on all-cause mortality. Hazard ratios ranged from 0.51 to 1.75. Of all specifications, 435 (36.0%) yielded hazard ratios equal to or above 1.0 and 773 (64.0%) below 1.0.

**Figure 2:**
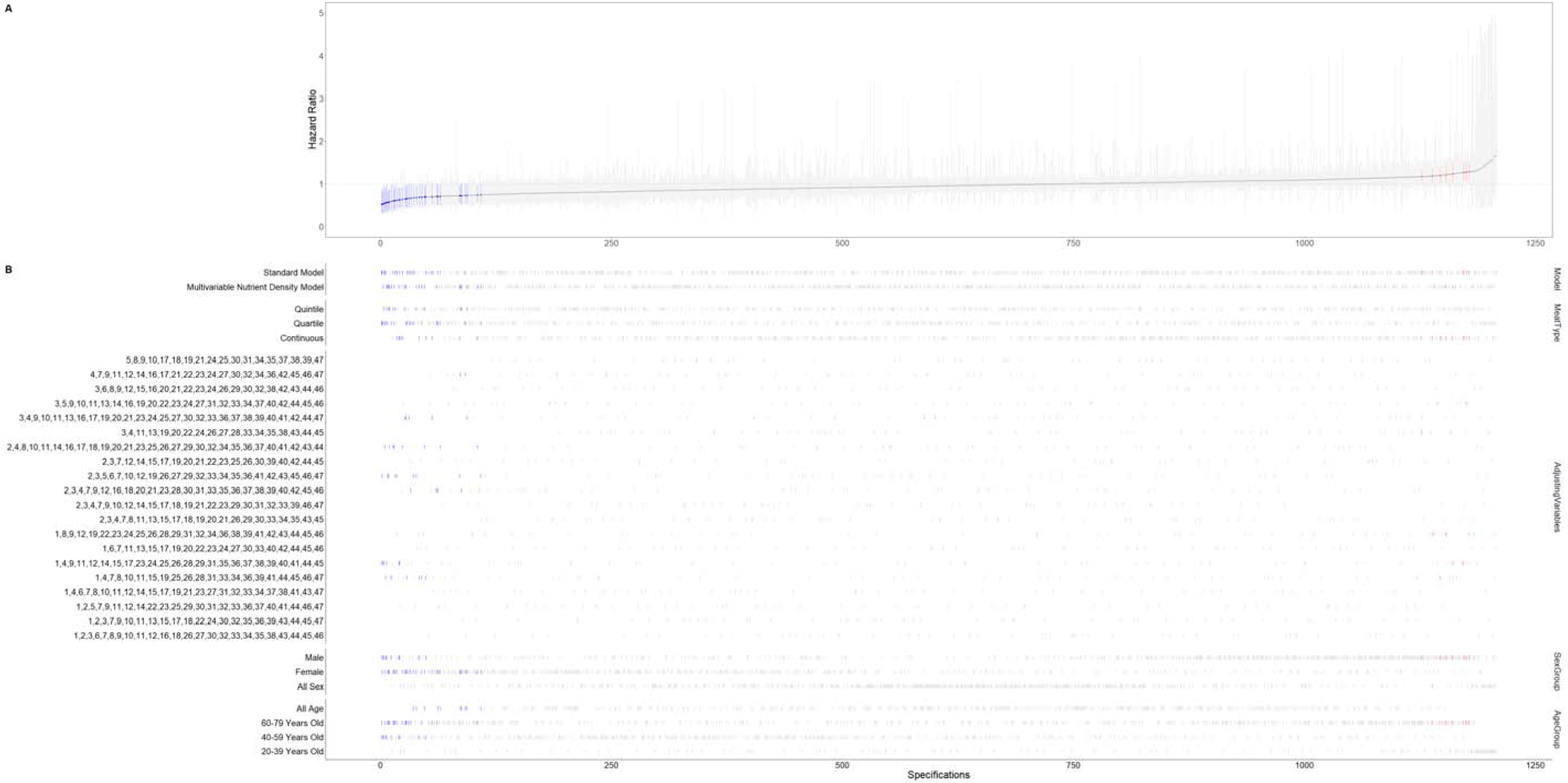
**Results of specification curve analysis.**

Of all specifications, 48 (3.97%) were statistically significant. Of 48 statistically significant results, 40 had significant point estimates that indicated red meat to reduce all-cause mortality and 8 indicated red meat to increase all-cause mortality. Among statistically significant effects suggesting benefit, we observed a median hazard ratio of 0.65 [IQR: 0.58 to 0.69] and, among statistically significant effects suggesting harm, we observed a median hazard ratio of 1.22 [IQR: 1.19 to 1.27]. We found 45% (542/1,208) of all specifications to yield point estimates ranging between HR of 0.90 to 1.10.

Visual inspection of the specification curve plot suggests subgroup by sex to importantly influence results, with analyses restricted to women more likely to suggest red meat is beneficial. We observed a median hazard ratio of 1.05 [IQR: 0.89 to 1.12] for men and 0.85 [IQR: 0.77 to 0.93] for women. We did not identify other analytic characteristics as consequential.

Supplement Figure 2 presents the results of the specification curve analysis stratified by how red meat is defined in analytic models (i.e., quartiles, quintiles, or continuous 100 g/day).

Supplement Tables 6 to 10 and Supplement Figures 3 to 7 show the results of tests for the proportional hazards assumption and graphical displays of the correlation between Schoenfeld residuals and ranked failure time. We did not find evidence that the proportional hazards assumption was violated in any analyses.

Finally, we present statistical inferences about the degree to which findings across all specifications are inconsistent with the null hypothesis (i.e., red meat has no effect on all-cause mortality) (Table 2). We performed statistical tests addressing whether the median effect estimate across all specifications is more extreme than expected if red meat had no effect on all-cause mortality, whether the proportion of specifications that produced statistically significant effects is more extreme than would expected if red meat had no effect on all-cause mortality, and whether Stouffer’s averaged Z value across all specifications is more extreme than would be expected if red meat had no effect on all-cause mortality. All three statistical tests yielded P-values > 0.05.

**Table 2:** Inferential statistics.

| Test statistics used | Observed results | P value<br>(% of bootstrap sample with results as or more extreme) |
| --- | --- | --- |
| Median effect size | HR=0.94 | P=0.472 |
| Share of significant results | 48 of 1208 specifications | P=0.998 |
| Aggregate all P values | Stouffer Z=-11.69 | P=0.732 |

## Discussion

### Main findings

In this study, we applied specification curve analysis—a method that involves defining and implementing all plausible and valid analytic approaches—to estimate the effect of unprocessed red meat on all-cause mortality (27). To mitigate the subjectivity involved in selecting analytic specifications, we sourced analytic approaches from the literature (29). We performed 1,208 unique analyses and found considerable variability in results, with hazard ratios ranging from 0.51 to 1.75. Our results suggest that findings in nutritional epidemiology studies may be contingent on analytic methods.

In contrast to previous studies addressing red meat, we found few of our analytic specifications to yield statistically significant effects. This may be because we used more recent data from NHANES, which includes fewer accumulated deaths (40). The most recent iterations of NHANES, however, are likely more reflective of the effects of red meat on all-cause mortality in the context of contemporaneous diets and lifestyles. Nevertheless, our primary objective was not draw inferences about the health effects of red meat but to provide a proof-of-concept illustration of the application of specification curve analysis to nutritional epidemiology.

Concerns may arise over the impact of various analytical techniques on the interpretation of results. For example, different methods for energy adjustment may have different implications for how the effect is interpreted (18, 41). In our study, we show that despite differences in analytic methods authors stated similar objectives and similarly interpreted their results. This suggests that authors are using disparate analytic methods to investigate near identical causal questions.

In addition to analytic flexibility, researchers criticize observational nutritional epidemiology studies for producing unreliable results due to biases associated with self-reported dietary data (20, 23). Nevertheless, nutritional epidemiology studies continue to play a critical role in shaping dietary recommendations and policies (15). While specification curve analysis does not address biases due to dietary measures, when combined with other tools and methods for producing more reliable dietary measures, specification curve analysis may have the potential to enhance confidence in the discipline (42, 43).

### Relation to previous work

Current evidence shows that results from studies may vary due to alternative analytic specifications and that there is often limited consensus on the optimal approach for data analysis (6, 44). Research to date has not, however, quantified the magnitude of variation in results for typical epidemiologic questions.

Further, while specification curve analysis has been previously applied in psychology and economics, it has not yet been applied in epidemiology or nutritional epidemiology (26, 45, 46).

### Strengths and limitations

The current work offers an innovative solution to analytic flexibility in nutritional epidemiology. To our knowledge, our work is the first application of specification curve analysis to nutritional epidemiology.

Our study also has limitations. There may be disagreements among investigators about what constitutes a justifiable analytical approach. To mitigate this issue, our choice of analytic specifications was informed by primary studies and so represents real, published analyses rather than possible, unpublished analyses that may only be marginally defensible. Further, we confirmed that the research questions addressed in the primary studies were adequately similar by collecting data on the objectives of primary studies and the ways in which authors interpreted their results.

We emphasize that specification curve analysis does not eliminate the need for subjectivity in selecting specifications (27). Nonetheless, it does improve on current practice in which investigators can test many alternative analytic specifications and selectively report results for those that yield interesting or favorable results. Specification curve analysis can identify findings that are most robust to alternative analytic specifications and encourage evidence users to interpret the results of nutritional epidemiology studies considering the typical variation in results expected due to analytic flexibility.

Different analytic methods may have implications for how the results are interpreted. For example, different methods to adjust for energy intake in nutritional epidemiology address different causal questions (18). Authors of nutritional epidemiology studies, however, seldom acknowledge these issues. We show that despite differences in analytic methods authors stated similar objectives and similarly interpreted their results.

Specification curve analysis also does not eliminate the need for content knowledge and expertise. Content knowledge and expertise are essential for selecting justifiable analytic specifications and interpreting the results of analyses.

We only applied specification curve analysis to one question—the effect of red meat on all-cause mortality. The extent to which results may be contingent on analytic methods may be different for other questions. We acknowledge that this is a controversial question in the nutrition literature and that the application of specification curve analytic to less contentious questions in nutritional epidemiology may improve its adoption. Our choice of topic was influenced by our team’s familiarity with red meat and the related literature (15, 29).

Our study likely underestimates the variations in results due to alternative analytic specifications since the analytic specifications that we could implement were limited by the availability of variables and data in NHANES. For example, due to the cross-sectional nature of NHANES, we were unable to use time-varying covariates and explore how alternative ways to account for these variables may influence results. There are also subjective analytic decisions in translating dietary recalls to nutrient and food intake, though we could not account for these decisions. For example, nutritional epidemiologists code dietary recalls according to food classification systems and subsequently use nutrition databases to estimate individual nutrient components of each item in dietary recalls—all of which involves subjective decisions.

Likewise, the continuous 2007-2014 NHANES data is likely suboptimal for investigating the effect of red meat and other nutritional exposures on health outcomes, due to it including few deaths and only collecting data on diet at a single point in time (30, 32). Nevertheless, our primary objective is not to provide conclusive answers about the health effects of red meat but to demonstrate a proof-of-concept application of specification curve analysis to nutritional epidemiology.

We did not incorporate weights in our analytic models. Sample weights in NHANES are designed to account for oversampling of specific subgroups and unequal probabilities of selection in the population. These weights are essential when the objective is to make inferences about population characteristics or to estimate prevalence rates because they adjust for factors that influence these estimates and ensure that the results are representative of the target population. However, when focusing on causal inference, the primary concern is to eliminate or control for confounding factors that may distort the true relationship between exposure and outcome and sample weights are less important, especially when variables used to derive sample weights are already included in analytic models (47–49).

We excluded results from models that yielded results that we deemed to be implausible based on pragmatic but arbitrary thresholds (i.e., HR ≤0.2 or HR ≥5). We suspect that the observed implausible specifications were due to sparse data bias—where there are too few events in critical combinations of explanatory variables (36). It is, however, possible that there were other models that produced results within this threshold that had too few events to reliably estimate the effect of red meat on all-cause mortality.

Finally, while we attempted to test the proportional hazards assumption using the correlation between Schoenfeld residuals and ranked failure time, these tests have limited sensitivity (50). We also only tested a proportion of our models for proportional hazards and it is possible that the proportional hazards assumption may be violated in models that we did not test.

### Implications

Specification curve analysis allows investigators to test all plausible and justifiable models to explain conflicting findings or contextualize emerging findings. While this study may provide insights on the health effects of unprocessed red meat, we believe the most important contribution of this study is to provide a proof-of-concept demonstrating the feasibility of applying specification curve analysis to nutritional epidemiology.

Nutritional epidemiology has long been criticized for producing sensational and conflicting findings, which has eroded confidence in the discipline (23). Nevertheless, nutritional epidemiology studies continue to play a crucial role in shaping dietary recommendations and policies, making it imperative to draw credible inferences from these studies (14, 15, 24). The broader application of specification curve analysis to nutritional epidemiology may enhance confidence in nutrition as a field by encouraging investigators to acknowledge an additional source of uncertainty in studies. When combined with other tools and methods that also address other limitations of observational nutritional epidemiology studies (e.g., biases that affect self-reported dietary data) (42), specification curve analysis has the potential to address a critical issue in epidemiology—analytic flexibility—and identify findings that are most robust to subjective analytic choices.

Findings from our study and future application of specification curve analysis will also be useful to evidence users who can interpret results in nutritional epidemiology studies in the context of the typical variation in results expected due to analytic flexibility. When effect estimates exceed the typical variation due to analytic methods, evidence users can be more certain of the findings, since they are likely robust to alternative analytic decisions.

Our findings may also have implications for precision nutrition that attempts to distinguish between subgroups of individuals who may differently respond to nutritional interventions or have different nutritional needs (51–53). Investigators have raised concerns that efforts to identify “responders” and realize precision nutrition may be highly dependent on the characteristics of analytic models (54).

Specification curve analysis may be useful for evaluating the reliability of precision nutrition claims across a range of defensible models.

We acknowledge that the application of specification curve analysis is time consuming and resource intensive. Sourcing justifiable analytic specifications from primary studies adds to this effort. While the application of specification curve analysis may not be feasible for all nutritional epidemiology questions, it can be applied to the most critical, impactful, or contentious questions in the discipline and can serve as an additional available tool to evaluate the credibility of nutrition claims in the literature.

## Conclusion

In this study, we apply specification curve analysis—a novel analytic method that involves defining and implementing all plausible and valid analytic approaches for addressing a research question—to investigate the effect of red meat on all-cause mortality. We show variability in results across plausible analytic specifications. Our study demonstrates the feasibility of applying specification curve analysis to nutritional epidemiology that offers a pragmatic and innovative solution to analytic flexibility. Specification curve analysis, in combination with other tools and methods, has the potential to improve the credibility of inferences from such studies.

This figure presents the results of the specification curve analysis, including 1,208 unique analytic specifications. The upper portion of the plot show HRs representing the effect of red meat on all-cause mortality. On the x-axis are the unique analytic specifications. The y-axis represents the magnitude of effect estimates. Each point on the graph represents the results of a unique analytic specification. Point estimates are shown in dark grey and 95% confidence intervals as light grey bars. Each point represents the results for the effect of red meat on all-cause mortality for a unique model. Points in blue are statistically significant and suggest red meat to prevent all-cause mortality and points in red are statistically significant and indicate red meat to increase risk of all-cause mortality.

The lower part of the plot show the characteristics of each analysis, including type of analytic model, operationalizations of variables, choice of covariates, and subgroups of interest. Each vertical line denotes the specific choice applied for each aspect of the analysis. We assigned a unique number to each covariate (Supplement 4 shows the number corresponding to each variable). Combinations of numbers in the graph represent combinations of covariates included in the model.

## Data Availability

Upon a reasonable request.

## Acknowledgments

None.

