## Supplement for "Variations in the results of nutritional epidemiology studies due to analytic flexibility: Application of specification curve analysis to red meat and all-cause mortality"

### Supplement Table 1: Study characteristics of primary studies examining unprocessed red meat or mixed unprocessed red meat and processed meat consumption and all-cause mortality.

| **Author** | **Year** | **Cohort(s)** | **Country** | **Participants (n)** | **Age at baseline** | **% Female** | **Duration of follow-up (years)** | **Exposures** |
| --- | --- | --- | --- | --- | --- | --- | --- | --- |
| S.M. Alshahrani | 2019 | Adventist Health Study 2 (AHS-2) | United States | 72,149 | Mean: 56.39 | 65.68 | Mean: 11.8 | Unprocessed red meat |
| R. Kappeler | 2013 | National Health and Nutrition Examination Survey III (NHANES III) | United States | 17,611 | Mean: 41.42 | 53.22 | Up to: 22 | Total red meat |
| L.J. Dominguez | 2018 | Seguimiento Universidad de Navarra (SUN) | Spain | 18,540 | NR | NR | Mean: 9.5 | Total red meat |
| A. Bellavia | 2016 | Cohort of Swedish Men (COSM), Swedish Mammography Cohort (SMC) | Sweden | 74,645 | Mean: 60.28 | 46.29 | Up to: 16 | Total red meat |
| A.Pan | 2012 | Health Professionals Follow-up Study (HPFS) | United States | 37,698 | Mean: 52.72 | 0 | Up to: 22 | Unprocessed red meat |
| A.Pan | 2012 | Nurses' Health Study (NHS) | United States | 83,644 | Mean: 46.08 | 100 | Up to: 28 | Unprocessed red meat |
| S. Rohrmann | 2013 | European Prospective Investigation into Cancer and Nutrition (EPIC) | France, Italy, Spain, The Netherlands, United Kingdom, Greece, Germany, Sweden, Norway, Denmark | 448,568 | Median: 52.3 (men); 50.9 (women) | 71.62 | Median: 12.7 | Total red meat |
| Y. Takata | 2013 | Shanghai Men's Health Study (SMHS) | China | 61,142 | Mean: 55.45 | 0 | Median: 5.5 | Total red meat |
| Y. Takata | 2013 | Shanghai Women's Health Study (SWHS) | China | 73,167 | Mean: 52.85 | 100 | Median: 11.2 | Total red meat |
| J. E. Lee | 2013 | Cardiovascular Diseases Risk Factor Two-Township Study (CVDFACTS) | Taiwan | 3,472 | Range: 18 to 92 | 56.45 | Mean: 14.9 (men), 15.6 (women) | Total red meat |
| J. E. Lee | 2013 | Health Effects of Arsenic Longitudinal Study (HEALS) | Bangladesh | 11,396 | Range: 17 to 75 | 57.14 | Mean: 6.6 | Total red meat |
| J. E. Lee | 2013 | Japan Public Health Center-based Prospective study cohort I (JPHC I) | Japan | 43,038 | Range: 40 to 59 | 52.15 | Up to: 14.2 (men), 14.7 (women) | Total red meat |
| J. E. Lee | 2013 | Japan Public Health Center-based Prospective study cohort II (JPHC II) | Japan | 56,411 | Range: 40 to 69 | 52.63 | Up to: 11.2 (men), 11.7 (women) | Total red meat |
| J. E. Lee | 2013 | Miyagi Cohort Study (Miyagi) | Japan | 44,966 | Range: 40 to 64 | 52.11 | Mean: 12.6 (men), 12.9 (women) | Total red meat |
| J. E. Lee | 2013 | Ohsaki National Health Insurance Cohort Study (Ohsaki) | Japan | 48,905 | Range: 40 to 80 | 52.03 | Mean: 9.8 (men), 10 (women) | Total red meat |
| J. E. Lee | 2013 | Seoul Male Cohort Study (Seoul) | South Korea | 13,600 | Range: 25 to 82 | 0 | Mean: 14.7 | Total red meat |
| J. E. Lee | 2013 | Shanghai Women's Health Study (SWHS) | China | 74,933 | Range: 40 to 71 | 100 | Mean: 8.6 | Total red meat |
| A. Etemadi | 2017 | National Institute of Health-American Association of Retired Persons (NIH-AARP) Diet and Health Study | United States | 536,969 | Mean: 62.16 | 41.06 | Median: 15.6 | Unprocessed red meat |
| M. S. Farvid | 2017 | Golestan Cohort Study | Iran | 42,466 | Mean: 51.62 | 56.93 | Median: 8.1 | Total red meat |
| D. A. Boggs | 2015 | Black Women's Health Study (BWHS) | United States | 37,001 | Mean: 41.89 | 100 | Up to: 16 | Red or processed meat |
| L. E. Kelemen | 2005 | Iowa Women's Health Study (IWHS) | United States | 29,017 | Median: 76.2 (Q1); 76.0 (Q2); 75.9 (Q3); 75.7 (Q4); 75.4 (Q5) | 100 | Up to: 15 | Total red meat |
| L. M. Nilsson | 2012 | Västerbotten Intervention Program (VIP) Cohort | Sweden | 77,319 | Median: 49 | 51.32 | Median: 10 | Red meat |
| D. Whiteman | 1999 | OXCHECK Study | United Kingdom | 10,522 | Range: 35 to 64 | 53.16 | Up to: 9 | Other fresh or frozen red meat |
| G. E. Fraser | 1999 | Adventist Health Study | United States | 34,198 | Mean: 54.23 | 59.48 | Up to: 6 | Meat eaters compared with vegetarians |

### Supplement Table 2: Analytical choices (regression, variable type of red meat, nutrition model, subgroup analysis) used in primary studies.

| Author | Regression | Exposure | Operationalization of red meat | Nutrition model used | Subgroup analysis | Any correction for measurement error | Number of unique models * | Substitutions | Exclusions | Missing data | Time-varying variables | Objective | Interpretation |
| --- | --- | --- | --- | --- | --- | --- | --- | --- | --- | --- | --- | --- | --- |
| S.M. Alshahrani | Time-dependent Cox regression | Unprocessed red meat | continuous (log-transformed) comparing 90th percentile with 0, 5 categories (zero intake group plus quartiles of consumers) | Residual model | Sex, race | Yes, only for exposure of interest | 9 | No | missing dietary variables, implausible responses to questionnaires, Canadian residents, extreme values of total energy intake (i.e., less than 500 kcal/day or more than 4500 kcal/day), younger than 25 years, prevalent cancer, cardiovascular disease | Multiple imputation | No | The Adventist Health Study-2 (AHS-2) has a large number of vegetarians, with even most nonvegetarians having low intakes of red and processed meat [10]. Therefore, this analysis aims to investigate the association of such low intakes of red and processed meat with all-cause, CVD, and cancer mortality. | In the Adventist Health Study-2 (AHS-2), we found relatively low levels of consumption of red and processed meat to be positively associated with all-cause and CVD mortality in multivariable-adjusted models, compared to zero-intake. |
| R. Kappeler | Time-dependent Cox regression | Red meat | 5 categories with cut points | Unadjusted model | Sex | No | 2 | No | history of myocardial infarction, stroke, heart failure or cancer | Complete case | No | To enhance the evidence, we analyzed data from the Third National Health and Nutrition Examination Survey (NHANES III), a large, nationally representative study of the US population to investigate whether red, processed and white meat consumption increase the risk for total mortality as well as cause-specific mortality, including cancer and CVD mortality. | In this large cohort study, we observed that consumption of red meat, processed meat, white meat and fish was not significantly associated with increased or decreased risk of total mortality, cancer mortality and CVD mortality. |
| L.J. Dominguez | Time-dependent Cox regression | Red meat | Continuous, or 4 categories with cut points | Unadjusted model, Residual model | Sex, age | No | 15 | Yes | extreme values of total energy intake (i.e., less than 800 kcal/day or more than 6000 kcal/day for men and 500 kcal/day for women or more than 5500 kcal/day for women) | Complete case | No | In light of recent research in this area and of the possible implications for dietary recommendations, we aimed to prospectively assess both, the associations of meat consumption (red, processed, red þ processed, and total) as well as of SFA intake with the risk of death in the SUN (“Seguimiento Universidad de Navarra”) longitudinal study and to asses if age was an effect modifier of this association. | High consumption of red, total, and red þ processed meat was significantly, independently, and strongly associated with increased all-cause mortality among highly educated persons older than 45 years compared with those with low consumption. |
| A. Bellavia | Time-dependent Cox regression | Red meat | 5 categories with cut points | Standard model | Sex, fruit, and vegetable intake | No | 4 | No | history of cardiovascular disease or cancer, extreme energy intake (3 SDs from log-transformed mean), extreme red meat intake (more than 300 g/day) | Complete case | No | We therefore evaluated whether the association between red meat consumption and the risk of all-cause, CVD, and cancer-specific mortality differed across FV amounts in a large prospective cohort of Swedish men and women | We found that the shape of the dose-response associations between red meat and all investigated outcomes were consistent over all FV concentrations. Higher intakes of total red meat intake were progressively associated with an increased risk of overall and CVD mortality, and these negative associations were largely reduced when restricting the analyses to nonprocessed red meat. |
| A.Pan | Time-dependent Cox regression | Unprocessed red meat | Continuous, quintiles | Standard model, nutrient density model | Study cohorts | No | 10 | Yes | history of cardiovascular disease or cancer, extreme energy intake (i.e., less than 500 kcal/day or more than 3500 kcal/day) | Complete case | Yes | Therefore, we investigated the association between red meat intake and total and cause specific mortality in two large cohorts with repeated measures of diet and up to 28 years of follow-up: the Health Professionals Follow-up Study (HPFS) and Nurses' Health Study (NHS). We also estimated the associations of substituting other healthy protein sources for red meat with total and cause-specific mortality. | In these two large prospective cohorts of U.S. men and women, we found that a higher intake of red meat was associated with a significantly elevated risk of total, CVD and cancer mortality, and this association was observed for both unprocessed and processed red meat, with a relatively greater risk for processed red meat. Substitution of fish, poultry, nuts, legumes, low-fat dairy products, and whole grains for red meat was associated with a significantly lower risk of mortality. |
| S. Rohrmann | Time-dependent Cox regression | Red meat | Continuous, 6 categories with cut points | Unadjusted model, standard model | Sex, smoking, alcohol consumption, fruit and vegetable intake | Yes | 3 | No | extreme values of total energy intake (i.e., top and bottom 1%), cancer, stroke or myocardial infarction at baseline | Complete case | No | Within the European Prospective Investigation into Cancer and Nutrition (EPIC) including more than 500,000 participants from ten European countries and, thus, reflecting a very heterogeneous diet, we examined the association between meat consumption and the risk for overall and cause-specific mortality. | In the EPIC cohort, a high consumption of processed meat was related to moderately higher all-cause mortality. After correction for measurement error, red meat intake was no longer associated with mortality, and there was no association with the consumption of poultry. |
| Y. Takata | Time-dependent Cox regression | Red meat | Quintiles | Standard model | Sex/cohort | No | 2 | No | history of cancer, extreme values of total energy intake (i.e., less than 500 kcal/day or more than 4000 kcal/day), participants who died during the first year of observation | Complete case for missing outcome data, replaced missing covariates with most commonly occurring category | No | In this report, we investigated the association of red meat and poultry intakes with the risk of mortality from all causes and specific causes, including cancer and CVD, by using data from two prospective cohort studies of 134,290 adult women and men in Shanghai, China. | In our two large, population-based, cohort studies involving 134,290 middle-aged and elderly women and men in Shanghai, China, we found that red meat intake was positively associated with total mortality among men, but not among women. |
| J. E. Lee | Time-dependent Cox regression | Red meat | Quartiles | Standard model | Sex, education, cohort | No | 3 | No | NR | Complete case | No | We initiated the Asia Cohort Consortium (ACC) to understand chronic disease etiology in Asia, where the association between dietary factors and chronic diseases has not been extensively studied. In the current study, we 1) compared ecological trends in meat intake over the past 37 y between Asia and the United States and 2) assessed whether meat intake was associated with all-cause, cancer and CVD mortality by using individual, prospective data from a pooled analysis of Asian cohort studies involving 296,721 men and women | Our pooled analysis of 8 Asian prospective cohort studies did not provide evidence of a higher risk of mortality for total meat intake. For red meat and poultry, inverse associations were observed for mortality in both men and women. We found that fish intake was inversely associated with risk of mortality in women. No variation in the associations between mortality and total meat intake were observed by educational levels or study period, |
| A. Etemadi | Time-dependent Cox regression | Unprocessed red meat | Continuous, quintiles | Residual model, nutrient density model | Sex, age groups, smoking, socioeconomic status, BMI, alcohol drinking, general health status, etc. | Yes | 3 | Yes | extreme values of total energy intake; prevalent cancer before the study | Complete case | No | To determine the association of different types of meat intake and meat associated compounds with overall and cause specific mortality. | Our results show an increased risk of all cause mortality and death due to nine different causes associated with both processed and unprocessed red meat. |
| M. S. Farvid | Time-dependent Cox regression | Total red meat | Continuous, quintiles | Unadjusted model, standard model | Sex, smoking, wealth score | No | 5 | Yes | extreme values of total energy intake (i.e., less than 600 kcal/day or more than 4,200 kcal/day), history of cancer, diabetes, coronary heart disease, or stroke | Complete case for outcome, replacement with median for covariates | No | Therefore, to examine the association between red meat and mortality and to show the effect of other protein sources on mortality, the association of red meat, poultry, fish, eggs, and legumes consumption with mortality risk from all and specific causes was investigated, using data from a prospective cohort study of adult men and women in Golestan, Iran. | Furthermore, high red meat intake was associated with higher mortality risk among current smokers as well as participants with higher SES. Poultry, fish, and legume consumption was not related to reduced CVD or all-cause mortality risk among all participants. |
| D. A. Boggs | Time-dependent Cox regression | Red or processed meat | Quintiles | Standard model | None | No | 1 | No | younger than 30 years, history of cancer (except nonmelanoma skin cancer), myocardial infarction, stroke, or diabetes, extreme values of total energy intake (i.e., less than 400 kcal/day or more than 3800 kcal/day), pregnant at baseline, implausible BMI (less than 15 or more than 60 kg/m2) | Complete case for outcome, missing data for covariates were modeled as indicator variables | Yes | In the present report, we assess the association of dietary intake with all-cause and cause-specific mortality among African American women. | High intake of red and processed meat was associated with increased mortality rates (HR: 1.31; 95% CI: 1.08, 1.60 for highest vs. lowest quintiles; P-trend = 0.004), which explains the association observed for the Western dietary pattern (Table 2), given that red meat and processed meat were the foods with the highest factor loadings for this dietary pattern (15). |
| L. E. Kelemen | Time-dependent Cox regression | Red or processed meat | Quintiles | Nutrient density model | N/A | No | 1 | Yes | premenopausal, history of cancer other than skin cancer, diabetes, or heart disease, extreme values of total energy intake (i.e., less than 600 kcal/day or more than 5000 kcal/day) | Complete case | No | We investigated the relation of total protein substituted for carbohydrate, a change that would be expected while adhering to a high-protein diet, and the association of different protein sources substituted for one another with chronic disease and mortality using data collected from the Iowa Women’s Health Study. | In this prospective study of over 29,000 postmenopausal women followed for 15 years, we found inverse associations of vegetable protein and legume food sources and positive associations of dairy and red meat food sources for CHD mortality when substituted in place of carbohydrate. Red meat food sources were also associated with increased risk for all-cause mortality but not for any cancer outcome. |
| L. M. Nilsson | Time-dependent Cox regression | Red meat | 2 categories: >median or <median | Residual model | Sex | No | 1 | No | unrealistic food intake level values, defined as a ratio of total energy intake to estimated basal metabolic rate in the lowest 5th percentile or the highest 2.5th percentile, determined separately for sex, follow-up times shorter than 2 years | Complete case | No | Historical sources on the subject of Sami diet are scarce, which was the motivation behind our previous study, including interviews of elderly Sami and comparisons of dietary habits in present-day reindeer herding Sami, non-reindeer-herding Sami and non-Sami. Taken together, the historical and scientific literature suggest a diet characterised by high intakes of fatty fish, red meat (primarily reindeer), fat, blood and organ dishes, wild berries and boiled, unfiltered coffee, and low intakes of cultivated vegetables and fruit, bread, and fibre.  In order to explore possible health effects of a ‘‘traditional Sami’’ dietary pattern, we have, in a manner similar to the Mediterranean diet score, constructed and studied a traditional Sami diet score, as well as the individual components of the score, in relation to all- cause, cancer, and cardiovascular mortality. The setting was a large, mainly non-Sami population-based cohort in northern Sweden. In contrast to much research focusing on ethnic minorities, we thus examined a general population from a minority perspective. | In this largely non-Sami, population-based cohort study of 77,319 men and women in northern Sweden, with up to 19 years of follow up, higher traditional Sami diet scores were associated with a weak increase in all-cause mortality in men. |
| D. Whiteman | Time-dependent Cox regression | Unprocessed red meat | 3 categories with cut points | Unadjusted model | Participation in OXCHECK, long-standing disease, exercise | No | 10 | No | history of myocardial infarction or angina | Complete case | No | Few reports have been published relating disease incidence or mortality among industrialized populations to consumption of broad food groups (e.g. vegetables, red meat, milk, fruit), yet most non nutritionists are likely to perceive their dietary intake in this way. To address the question of whether simple dietary assessments are associated with risk of death from specific causes, we analysed the mortality experience of a prospective study of more than 11 000 patients in a British primary care setting with 9 years’ follow-up. | The principal dietary factors associated with reduced mortality risk in this study were frequent consumption of fresh or frozen green vegetables or salad, fresh or frozen red meat (such as beef, lamb or pork), and biscuits, cakes, puddings or sweets; consumption of these foods was most strongly associated with a reduced risk of IHD mortality. |
| G. E. Fraser ^‡^ | NR | Beef | 3 categories with cut points | Unadjusted model | Sex | No | 1 | No | NR | Complete case | No | For this report, we summarized findings associating the use of different foods to risk of cancer, ischemic heart disease (IHD), and other diseases within a Seventh-day Adventist population enrolled in a large cohort study (1976–1988) | Our findings strongly suggest that dietary factors have an important influence on longevity and the risk of a number of chronic diseases. |

*We considered a model unique if it used a different analytic method, method of adjustment for energy, operationalization of variables, combination of covariates, or subgroups, with two exceptions. We considered step wise models (where several models are presented each adjusting for the covariates included in the previous model and a set of additional covariates) as a single model. Furthermore, we considered subgroup analyses or stratified models (e.g., stratified based on sex) as a single model. When additive models were reported, we excluded substitution models. When studies only reported substitution models, we considered substitution models.

‡This article used 2 by 2 tables and reported Mantel-Haenszel pooled relative risk

### Supplement Table 3: Covariates considered in primary studies.

A green, white, grey box indicates variable adjusted/not adjusted/not applicable in the analysis for the corresponding primary study.

| **Author** | **S.M. Alshahrani** | **R. Kappeler** | **L.J. Dominguez** | **A. Bellavia** | **A.Pan** | **S. Rohrmann** | **Y. Takata** | **J. E. Lee** | **A. Etemadi** | **M. S. Farvid** | **D. A. Boggs** | **L. E. Kelemen** | **L. M. Nilsson** | **D. Whiteman** |  |
| --- | --- | --- | --- | --- | --- | --- | --- | --- | --- | --- | --- | --- | --- | --- | --- |
|  |  |  |  |  |  |  |  |  |  |  |  |  |  |  | **G. E. Fraser** |
| Age |  |  |  |  |  |  |  |  |  |  |  |  |  |  |  |
| Smoking |  |  |  |  |  |  |  |  |  |  |  |  |  |  |  |
| Alcohol use |  |  |  |  |  |  |  |  |  |  |  |  |  |  |  |
| Sex |  |  |  |  |  |  |  |  |  |  |  |  |  |  |  |
| Total energy intake |  |  |  |  |  |  |  |  |  |  |  |  |  |  |  |
| Physical activity |  |  |  |  |  |  |  |  |  |  |  |  |  |  |  |
| Vegetables |  |  |  |  |  |  |  |  |  |  |  |  |  |  |  |
| Fruits |  |  |  |  |  |  |  |  |  |  |  |  |  |  |  |
| Education |  |  |  |  |  |  |  |  |  |  |  |  |  |  |  |
| BMI |  |  |  |  |  |  |  |  |  |  |  |  |  |  |  |
| History of diabetes |  |  |  |  |  |  |  |  |  |  |  |  |  |  |  |
| History of hypertension |  |  |  |  |  |  |  |  |  |  |  |  |  |  |  |
| Seafood/fish |  |  |  |  |  |  |  |  |  |  |  |  |  |  |  |
| Poultry |  |  |  |  |  |  |  |  |  |  |  |  |  |  |  |
| Race/Ethnicity |  |  |  |  |  |  |  |  |  |  |  |  |  |  |  |
| Marital status |  |  |  |  |  |  |  |  |  |  |  |  |  |  |  |
| Legumes |  |  |  |  |  |  |  |  |  |  |  |  |  |  |  |
| Socioeconomic status |  |  |  |  |  |  |  |  |  |  |  |  |  |  |  |
| History of hypercholesterolemia |  |  |  |  |  |  |  |  |  |  |  |  |  |  |  |
| Processed meat |  |  |  |  |  |  |  |  |  |  |  |  |  |  |  |
| Use of hormone therapy |  |  |  |  |  |  |  |  |  |  |  |  |  |  |  |
| Eggs |  |  |  |  |  |  |  |  |  |  |  |  |  |  |  |
| Multivitamin use |  |  |  |  |  |  |  |  |  |  |  |  |  |  |  |
| Use of aspirin |  |  |  |  |  |  |  |  |  |  |  |  |  |  |  |
| History of cardiovascular disease |  |  |  |  |  |  |  |  |  |  |  |  |  |  |  |
| Whole grain |  |  |  |  |  |  |  |  |  |  |  |  |  |  |  |
| Family history of cancer |  |  |  |  |  |  |  |  |  |  |  |  |  |  |  |
| Nuts and seeds |  |  |  |  |  |  |  |  |  |  |  |  |  |  |  |
| Urban or rural residence |  |  |  |  |  |  |  |  |  |  |  |  |  |  |  |
| Sedentary lifestyle |  |  |  |  |  |  |  |  |  |  |  |  |  |  |  |
| Watching TV |  |  |  |  |  |  |  |  |  |  |  |  |  |  |  |
| History of cancer |  |  |  |  |  |  |  |  |  |  |  |  |  |  |  |
| History of stroke |  |  |  |  |  |  |  |  |  |  |  |  |  |  |  |
| Family history of diabetes |  |  |  |  |  |  |  |  |  |  |  |  |  |  |  |
| Use of statin |  |  |  |  |  |  |  |  |  |  |  |  |  |  |  |
| Total dairy |  |  |  |  |  |  |  |  |  |  |  |  |  |  |  |
| Dietary fiber |  |  |  |  |  |  |  |  |  |  |  |  |  |  |  |
| Coffee |  |  |  |  |  |  |  |  |  |  |  |  |  |  |  |
| Sugar-sweetened beverages |  |  |  |  |  |  |  |  |  |  |  |  |  |  |  |
| Menopausal status |  |  |  |  |  |  |  |  |  |  |  |  |  |  |  |
| Bread |  |  |  |  |  |  |  |  |  |  |  |  |  |  |  |
| Study center |  |  |  |  |  |  |  |  |  |  |  |  |  |  |  |
| Questionnaire cycle |  |  |  |  |  |  |  |  |  |  |  |  |  |  |  |
| Years of entering cohort |  |  |  |  |  |  |  |  |  |  |  |  |  |  |  |
| Occupation |  |  |  |  |  |  |  |  |  |  |  |  |  |  |  |
| Income |  |  |  |  |  |  |  |  |  |  |  |  |  |  |  |
| Sleep |  |  |  |  |  |  |  |  |  |  |  |  |  |  |  |
| Weight |  |  |  |  |  |  |  |  |  |  |  |  |  |  |  |
| Height |  |  |  |  |  |  |  |  |  |  |  |  |  |  |  |
| Systolic blood pressure |  |  |  |  |  |  |  |  |  |  |  |  |  |  |  |
| General health condition |  |  |  |  |  |  |  |  |  |  |  |  |  |  |  |
| Comorbidity index |  |  |  |  |  |  |  |  |  |  |  |  |  |  |  |
| History of depression |  |  |  |  |  |  |  |  |  |  |  |  |  |  |  |
| History of myocardial infarction |  |  |  |  |  |  |  |  |  |  |  |  |  |  |  |
| Family history of myocardial infarction |  |  |  |  |  |  |  |  |  |  |  |  |  |  |  |
| Family history of hypercholesterolemia |  |  |  |  |  |  |  |  |  |  |  |  |  |  |  |
| On special diet |  |  |  |  |  |  |  |  |  |  |  |  |  |  |  |
| Snacking between meals |  |  |  |  |  |  |  |  |  |  |  |  |  |  |  |
| Use of Ibuprofen |  |  |  |  |  |  |  |  |  |  |  |  |  |  |  |
| Use of opium |  |  |  |  |  |  |  |  |  |  |  |  |  |  |  |
| Use of Valsartan |  |  |  |  |  |  |  |  |  |  |  |  |  |  |  |
| Use of blood pressure medications |  |  |  |  |  |  |  |  |  |  |  |  |  |  |  |
| Low-fat dairy |  |  |  |  |  |  |  |  |  |  |  |  |  |  |  |
| Cheese |  |  |  |  |  |  |  |  |  |  |  |  |  |  |  |
| Cholesterol |  |  |  |  |  |  |  |  |  |  |  |  |  |  |  |
| SFA |  |  |  |  |  |  |  |  |  |  |  |  |  |  |  |
| MUFA |  |  |  |  |  |  |  |  |  |  |  |  |  |  |  |
| PUFA |  |  |  |  |  |  |  |  |  |  |  |  |  |  |  |
| TFA |  |  |  |  |  |  |  |  |  |  |  |  |  |  |  |
| Fatty fish |  |  |  |  |  |  |  |  |  |  |  |  |  |  |  |
| Fat |  |  |  |  |  |  |  |  |  |  |  |  |  |  |  |
| Berries |  |  |  |  |  |  |  |  |  |  |  |  |  |  |  |
| Blood dishes |  |  |  |  |  |  |  |  |  |  |  |  |  |  |  |
| Liver/kidney |  |  |  |  |  |  |  |  |  |  |  |  |  |  |  |
| Pudding |  |  |  |  |  |  |  |  |  |  |  |  |  |  |  |
| Cakes |  |  |  |  |  |  |  |  |  |  |  |  |  |  |  |
| Mediterranean diet score |  |  |  |  |  |  |  |  |  |  |  |  |  |  |  |
| Sodium |  |  |  |  |  |  |  |  |  |  |  |  |  |  |  |
| Oral contraceptive use |  |  |  |  |  |  |  |  |  |  |  |  |  |  |  |
| Dietary methionine |  |  |  |  |  |  |  |  |  |  |  |  |  |  |  |
| Vitamin E supplement |  |  |  |  |  |  |  |  |  |  |  |  |  |  |  |
| History of angina |  |  |  |  |  |  |  |  |  |  |  |  |  |  |  |
| Total grain |  |  |  |  |  |  |  |  |  |  |  |  |  |  |  |
| Soft drink or soda |  |  |  |  |  |  |  |  |  |  |  |  |  |  |  |
| Total meat |  |  |  |  |  |  |  |  |  |  |  |  |  |  |  |
| Magnesium |  |  |  |  |  |  |  |  |  |  |  |  |  |  |  |
| Carbohydrates |  |  |  |  |  |  |  |  |  |  |  |  |  |  |  |
| Biscuits and sweets |  |  |  |  |  |  |  |  |  |  |  |  |  |  |  |
| Parity |  |  |  |  |  |  |  |  |  |  |  |  |  |  |  |

### Supplement Table 4: Variables considered in specification curve analysis. Variables that have an index were treated as optional adjusting variables and each index in the specification curve plot corresponds to a variable.

| **Variable names** | **Variable type** | **Categories** | **Variable description** | **How variables treated in the analysis** | **Index in plot** |
| --- | --- | --- | --- | --- | --- |
| Years of entering cohort | Categorical | 2007-2008; 2009-2010; 2011-2012; 2013-2014 | Data Release Number. 4 categories. | Mandatory adjusting variable |  |
| Mortality status | Categorical | Assumed alive; Assumed deceased | The public-use LMF provides mortality follow-up data from the date of survey participation through December 31, 2019. The mortality status variable is the final determination of vital status and should be used as an outcome variable to calculate survival. 2 categories. | Outcome |  |
| Eligibility Status for Mortality Follow-up | Categorical | Eligible; Under age 18, not available for public release; Ineligible | The public-use LMF provides mortality follow-up data from the date of survey participation through December 31, 2019. Survey participants are defined as ineligible for mortality linkage if they had insufficient identifying data or are under the age of 18 and not available for public release. 3 categories. | Used to subset data |  |
| Number of Person Months of Follow-up from NHANES interview date | Continuous |  | The number of person-months of follow-up from the NHANES interview date. Participants who are assumed alive are assigned the number of person months at the end of the mortality period, December 31, 2019. | Outcome |  |
| Age | Continuous |  | Best age in years of the sample person at the time of HH screening. 20-79 years old considered. | Mandatory adjusting variable |  |
| Age groups | Categorical | 20-39 years old; 40-59 years old; 60-79 years old | Use the age in years to form 3 categories: 20-39 years old; 40-59 years old; 60-79 years old. | Used to subset data |  |
| Gender | Categorical | Male; Female | Gender of the sample person. 2 categories. | Mandatory adjusting variable used to subset data |  |
| Race/Ethnicity | Categorical | Mexican American; Other Hispanic; Non-Hispanic White; Non-Hispanic Black; Other Race - Including Multi-Racial | Reported race and ethnicity information. 5 categories. | Optional adjusting variable | 1 |
| Education Level | Categorical | Less Than 9th Grade; 9-11th Grade (Includes 12th grade with no diploma); High School Grad/GED or Equivalent; Some College or AA degree; College Graduate or above. | What is the highest grade or level of school completed or the highest degree received? 5 categories. | Optional adjusting variable | 2 |
| Marital Status | Categorical | Married; Widowed; Divorced; Separated; Never married; Living with a partner. | Marital Status of sample person. 6 categories. | Optional adjusting variable | 3 |
| Alcohol drinking | Continuous |  | In the past 12 months, on those days that you drank alcoholic beverages, on average, how many drinks did you have? | Optional adjusting variable | 4 |
| Alcohol drinking groups | Categorical | Non-drinker; Moderate drinker; Heavy drinker | Categorize alcoholic drinks per day into 3 categories: Non-drinker: =0; Moderate drinker: 0-2 drinks per day; Heavy drinker>=2 drinks per day. | Optional adjusting variable | 5 |
| Smoking | Categorical | Non or light smoker; Moderate smoker; Heavy smoker. | People who answered they never smoked cigarettes regularly or not smoked at least 100 cigarettes in life or smoked at least 100 cigarettes in life but quit are categorized as a non or light smoker. People who answered they smoked at least 100 cigarettes in life and continue smoking are categorized as follows: if >67 cigarettes per month (40 pack/year) then heavy smoker, if <=67 cigarettes per month (40 pack/year) then moderate smoker. For smoking, categorize as 3 categories: Non or light smoker; Moderate smoker; Heavy smoker. | Mandatory adjusting variable |  |
| Occupation | Categorical | Non-worker; Part-time worker; Full-time worker | How many hours did you work last week at all jobs or businesses? For occupation, we categorized participants into 3 categories: non-worker (0 hours a week); Part-time worker (1-30 hours a week); Full-time worker (>=31 hours a week). | Optional adjusting variable | 6 |
| Vigorous or moderate recreational activity | Categorical | Yes; No | The next questions exclude the work and transportation activities that you have already mentioned. Now I would like to ask you about sports, fitness and recreational activities. Do you do any vigorous-intensity sports, fitness, or recreational activities that cause large increases in breathing or heart rate like running or basketball for at least 10 minutes continuously? Do you do any moderate-intensity sports, fitness, or recreational activities that cause a small increase in breathing or heart rate such as brisk walking, bicycling, swimming, or golf for at least 10 minutes continuously? For physical activity, if the participants either do a vigorous recreational activity or a moderate recreational activity, then they are defined as they do physical activity. If they neither do a vigorous recreational activity nor a moderate recreational activity, then they are defined as don’t do physical activity. 2 categories. | Optional adjusting variable | 7 |
| Sedentary lifestyle | Categorical | Low; Lower-middle; Middle; Upper-middle; High | The following question is about sitting at work, at home, getting to and from places, or with friends, including time spent sitting at a desk, traveling in a car or bus, reading, playing cards, watching television, or using a computer. Do not include time spent sleeping. How much time do you usually spend sitting on a typical day? People whose sedentary lifestyle minutes answered >=1000 are treated as missing because they are probably including the time they sleep and that is not plausible. Categorize sedentary lifestyle as fifth. 5 categories. | Optional adjusting variable | 8 |
| Sleep | Categorical | ≤4 hours; 5-8 hours; ≥9 hours | The next set of questions is about your sleeping habits. How much sleep do you usually get at night on weekdays or workdays? Categorize sleep into 3 categories: <=4 hours, 5-8 hours, >=9 hours. | Optional adjusting variable | 9 |
| Annual family income | Categorical | $0-$14,999; $15000-$34999; $35000-$64999; Over $65000 | Total family income (reported as a range value in dollars). For Annual Family Income, create 4 categories: 1: $0-$14,999; 2: $15000-$34999; 3: $35000-$64999; 4: Over $65000. For Annual Family Income, subjects that answered over or under $20000 but not as a specific range are treated as missing. | Optional adjusting variable | 10 |
| Socioeconomic status | Categorical | Low; Lower-middle; Middle; Upper-middle; High | A ratio of family income to poverty threshold. We categorize the ratio of family income to poverty (PIR) into fifth to denote their socioeconomic status. 5 categories. | Optional adjusting variable | 11 |
| BMI | Continuous |  | Body Mass Index (kg/m^2). Exclude Implausible BMI (<15 or ≥60 kg/m^2) | Optional adjusting variable | 12 |
| BMI Group | Categorical | Healthy weight; Obesity; Overweight; Underweight | Categorize BMI into 4 categories: Underweight (<18.5), Healthy weight (18.5<=<25), Overweight (25<=<30), and Obesity (>=30), based on the definition by CDC. | Optional adjusting variable | 13 |
| Systolic blood pressure | Categorical | Low; Lower-middle; Middle; Upper-middle; High | Systolic: Blood pressure (1st, 2nd, 3rd, 4th reading) mm Hg. Systolic Blood pressure has 4 readings; we use the average of these four readings. Categorize systolic blood pressure into fifth. 5 categories. | Optional adjusting variable | 14 |
| Health condition | Categorical | Poor; Excellent; Very Good; Good; Fair | Would you say your health in general is . . . For general health conditions, categorize into 5 categories. | Optional adjusting variable | 15 |
| History of hypercholesterolemia | Categorical | Yes; No | Have you ever been told by a doctor or other health professional that your blood cholesterol level was high? 2 categories. | Optional adjusting variable | 16 |
| History of hypertension | Categorical | Yes; No | Have you ever been told by a doctor or other health professional that you had hypertension, also called high blood pressure? 2 categories. | Optional adjusting variable | 17 |
| History of diabetes | Categorical | Yes; No | The next questions are about specific medical conditions. Other than during pregnancy, have you ever been told by a doctor or health professional that you have diabetes or sugar diabetes? Borderline diabetes is treated as no diabetes. 2 categories. | Optional adjusting variable | 18 |
| History of depression | Categorical | Yes; No | Over the last 2 weeks, how often have you been bothered by the following problems: feeling down, depressed, or hopeless? For depression, we define a subject that answers several days or more than half the days or nearly every day as having a history of depression. 2 categories. | Optional adjusting variable | 19 |
| History of cardiovascular disease | Categorical | Yes; No | Has a doctor or other health professional ever told you that you had coronary heart disease? Has a doctor or other health professional ever told you you had a stroke? History of cardiovascular disease is defined as participants having a history of coronary heart disease or history of stroke. 2 categories. | Optional adjusting variable | 20 |
| History of cancer or malignancy | Categorical | Yes; No | Have you ever been told by a doctor or other health professional that you had cancer or a malignancy of any kind? 2 categories. | Optional adjusting variable | 21 |
| Family history of diabetes | Categorical | Yes; No | Including living and deceased, were any of your close biological that is, blood relatives including father, mother, sisters, or brothers, ever told by a health professional that they had diabetes? 2 categories. | Optional adjusting variable | 22 |
| Family history of myocardial infarction | Categorical | Yes; No | Including living and deceased, were any of your close biological that is, blood relatives including father, mother, sisters, or brothers, ever told by a health professional that they had a heart attack or angina (an-gi-na) before the age of 50? 2 categories. | Optional adjusting variable | 23 |
| Menopausal status (women) | Categorical | Premenopausal; Postmenopausal | Menopausal status: premenopausal, postmenopausal | Mandatory adjusting variable |  |
| Hormone therapy use (women) | Categorical | Yes; No | Have you ever used female hormones such as estrogen and progesterone? Please include any forms of female hormones, such as pills, creams, patches, and injectables, but do not include birth control methods or use for infertility. 2 categories. | Mandatory adjusting variable |  |
| Parity (women) | Categorical | Nulliparous; Parous | The next questions are about your pregnancy history. Have you ever been pregnant? Please include (current pregnancy,) live births, miscarriages, stillbirths, tubal pregnancies, and abortions. 2 categories. | Mandatory adjusting variable |  |
| Oral contraceptive use (women) | Categorical | Yes; No | Now I am going to ask you about your birth control history. Have you ever taken birth control pills for any reason? 2 categories. | Mandatory adjusting variable |  |
| Pregnant at baseline (women) | Categorical | Yes; No; . | Are you pregnant now? 3 categories. Exclude Women pregnant at baseline, women missing pregnant information are treated as non-pregnant at baseline. | Used to subset data |  |
| Use of Aspirin | Categorical | Yes; No | We searched records of prescription medication use and identified users of aspirin, atorvastatin, ibuprofen, opium, statin, and valsartan users. If we did not find any record of a medication for a person, then it is treated as a non-user. 2 categories. | Optional adjusting variable | 24 |
| Use of Ibuprofen | Categorical | Yes; No | We searched records of prescription medication use and identified users of aspirin, atorvastatin, ibuprofen, opium, statin, and valsartan users. If we did not find any record of a medication for a person, then it is treated as a non-user. 2 categories. | Optional adjusting variable | 25 |
| Use of Opium | Categorical | Yes; No | We searched records of prescription medication use and identified users of aspirin, atorvastatin, ibuprofen, opium, statin, and valsartan users. If we did not find any record of a medication for a person, then it is treated as a non-user. 2 categories. | Optional adjusting variable | 26 |
| Use of Statin | Categorical | Yes; No | We searched records of prescription medication use and identified users of aspirin, atorvastatin, ibuprofen, opium, statin, and valsartan users. If we did not find any record of a medication for a person, then it is treated as a non-user. 2 categories. | Optional adjusting variable | 27 |
| Use of Valsartan | Categorical | Yes; No | We searched records of prescription medication use and identified users of aspirin, atorvastatin, ibuprofen, opium, statin, and valsartan users. If we did not find any record of a medication for a person, then it is treated as a non-user. 2 categories. | Optional adjusting variable | 28 |
| Special diet | Categorical | Yes; No | Are you currently on any kind of diet, either to lose weight or for some other health-related reason? 2 categories. | Optional adjusting variable | 29 |
| Use of dietary supplement | Categorical | Yes; No | Any Dietary Supplements taken in the past 24 hours? 2 categories. | Optional adjusting variable | 30 |
| Processed meat | Continuous |  | For individual food components, we averaged day 1 and day 2. Processed meat is defined as curved meat: Frankfurters, sausages, corned beef, and luncheon meat that are made from beef, pork, or poultry. gram eq. used. | Optional adjusting variable | 31 |
| Unprocessed red meat | Continuous |  | For individual food components, we averaged day 1 and day 2. Unprocessed red meat is defined as Beef, veal, pork, lamb, and game meat; excludes organ meat and cured meat. gram eq. used. | Exposure variable |  |
| Poultry | Continuous |  | For individual food components, we averaged day 1 and day 2. Poultry is defined as Chicken, turkey, Cornish hens, duck, goose, quail, and pheasant (game birds); excludes organ meat and cured meat. gram eq. used. | Optional adjusting variable | 32 |
| Fruits | Continuous |  | For individual food components, we averaged day 1 and day 2. If both are missing, then missing. Fruit is defined as Total intact fruits (whole or cut) but excludes fruit juices. Cup eq. used. | Optional adjusting variable | 33 |
| Vegetables | Continuous |  | For individual food components, we averaged day 1 and day 2. Vegetables include Total dark green, red and orange, starchy, and other vegetables; excludes legumes. Cup eq. used. | Optional adjusting variable | 34 |
| Seafood | Continuous |  | For individual food components, we averaged day 1 and day 2. Seafood is defined as Seafood (finfish, shellfish, and other seafood) high in n-3 fatty acids+ Seafood (finfish, shellfish, and other seafood) low in n-3 fatty acids. gram eq. used. | Optional adjusting variable | 35 |
| Whole grain | Continuous |  | For individual food components, we averaged day 1 and day 2. Grains are defined as whole grains and contain the entire grain kernel ― the bran, germ, and endosperm. gram eq. used. | Optional adjusting variable | 36 |
| Eggs | Continuous |  | For individual food components, we averaged day 1 and day 2. Eggs are defined as Eggs (chicken, duck, goose, quail) and egg substitutes. gram eq. used. | Optional adjusting variable | 37 |
| Nuts and seeds | Continuous |  | For individual food components, we averaged day 1 and day 2. Nuts and seeds are defined as Peanuts, tree nuts, and seeds; excludes coconut. gram eq. used. | Optional adjusting variable | 38 |
| Legumes | Continuous |  | For individual food components, we averaged day 1 and day 2. Legumes are defined as Beans and Peas. gram eq. used. | Optional adjusting variable | 39 |
| Total dairy | Continuous |  | For individual food components, we averaged day 1 and day 2. Total diary is defined as Milk type+ Yogurt type + Cheese type. Cup eq. used. | Optional adjusting variable | 40 |
| Total energy | Continuous |  | For total nutrition intake variables (TCAL, TCARB ….), we averaged day 1 and day 2. Energy (kcal/d). Exclude Extreme value of total energy intake (<500kcal/d or >4500kcal/d) | Mandatory adjusting variable |  |
| Carbohydrates | Continuous |  | For total nutrition intake variables (TCAL, TCARB ….), we averaged day 1 and day 2. Carbohydrate (gm/d) | Optional adjusting variable | 41 |
| Dietary fiber | Continuous |  | For total nutrition intake variables (TCAL, TCARB ….), we averaged day 1 and day 2. Dietary fiber (gm/d) | Optional adjusting variable | 42 |
| Saturated fatty acid | Continuous |  | For total nutrition intake variables (TCAL, TCARB ….), we averaged day 1 and day 2. Total saturated fatty acids (gm/d) | Optional adjusting variable | 43 |
| Monounsaturated fatty acid | Continuous |  | For total nutrition intake variables (TCAL, TCARB ….), we averaged day 1 and day 2. Total monounsaturated fatty acids (gm/d) | Optional adjusting variable | 44 |
| Polyunsaturated fatty acid | Continuous |  | For total nutrition intake variables (TCAL, TCARB ….), we averaged day 1 and day 2. Total polyunsaturated fatty acids (gm/d) | Optional adjusting variable | 45 |
| Cholesterol | Continuous |  | For total nutrition intake variables (TCAL, TCARB ….), we averaged day 1 and day 2. Cholesterol (mg/d) | Optional adjusting variable | 46 |
| Magnesium | Continuous |  | For total nutrition intake variables (TCAL, TCARB ….), we averaged day 1 and day 2. Magnesium (mg/d) | Optional adjusting variable | 47 |

### Supplement Table 5: Characteristics of participants included in specification curve analyses from continuous NHANES 2007-2014

| **All Included Variables*** | | | | |
| --- | --- | --- | --- | --- |
| **Total participants**, N | 10661 |  | **Years of entering cohort** |  |
| **All-Cause mortality**, n (%) | 1022 (10) |  | 2007-2008, n (%) | 2311 (22) |
| **Months follow-up** (months) | 99 (65, 143) |  | 2009-2010, n (%) | 2358 (22) |
|  |  |  | 2011-2012, n (%) | 2857 (27) |
| **Age** (years) | 50 (27, 71) |  | 2013-2014, n (%) | 3135 (29) |
| **Age groups** |  |  |  |  |
| 20-39 years old, n (%) | 3210 (30) |  | **Reproductive health for women** |  |
| 40-59 years old, n (%) | 3985 (37) |  | Postmenopausal, n (%) | 2653 (52) |
| 60-79 years old, n (%) | 3466 (33) |  | Use of hormone therapy, n (%) | 1186 (23) |
| **Gender** |  |  | Parous women, n (%) | 4360 (85) |
| Female, n (%) | 5150 (48) |  | Oral contraceptive use, n (%) | 3940 (77) |
| Male, n (%) | 5511 (52) |  |  |  |
|  |  |  | **Smoking** |  |
| **Dietary Intakes** |  |  | Non or light smoker, n (%) | 8373 (79) |
| Unprocessed red meat (g/d) | 29.5 (0, 120.2) |  | Moderate smoker, n (%) | 437 (4) |
| Total energy intake (kcal/d) | 1945 (1168, 3099) |  | Heavy smoker, n (%) | 1851 (17) |
| **Optional adjusting variables*** | | | | |
| **(1) Race/Ethnicity** |  |  | **(14) Systolic blood pressure** (mm Hg) ^†^ | 120.7 (104.0, 145.3) |
| Mexican American, n (%) | 1321 (12) |  | **(15) Health condition** |  |
| Other Hispanic, n (%) | 988 (9) |  | Poor, n (%) | 395 (4) |
| Non-Hispanic White, n (%) | 5193 (49) |  | Excellent, n (%) | 976 (9) |
| Non-Hispanic Black, n (%) | 2235 (21) |  | Very good, n (%) | 3105 (29) |
| Other Race - Including Multi-Racial, n (%) | 924 (9) |  | Good, n (%) | 4226 (40) |
| **(2) Education** |  |  | Fair, n (%) | 1959 (18) |
| Less Than 9th Grade, n (%) | 719 (7) |  | **History of diseases** |  |
| 9-11th Grade (Incl. 12th w/o diploma), n (%) | 1443 (14) |  | (16) Hypercholesterolemia, n (%) | 4193 (39) |
| High School Grad/GED or Equivalent, n (%) | 2297 (22) |  | (17) Hypertension, n (%) | 4086 (38) |
| Some College or AA degree, n (%) | 3336 (31) |  | (18) Diabetes, n (%) | 1378 (13) |
| College Graduate or above, n (%) | 2866 (27) |  | (19) Depression, n (%) | 2576 (24) |
| **(3) Marital status** |  |  | (20) Cardiovascular disease, n (%) | 713 (7) |
| Never married, n (%) | 1726 (16) |  | (21) Cancer or malignancy, n (%) | 1041 (10) |
| Married, n (%) | 5891 (55) |  | (22) Family history of diabetes, n (%) | 4510 (42) |
| Widowed, n (%) | 560 (5) |  | (23) Family history of myocardial infarction, n (%) | 1395 (13) |
| Divorced, n (%) | 1366 (13) |  | **Prescription medication intakes** |  |
| Separated, n (%) | 339 (3) |  | (24) Use of Aspirin, n (%) | 172 (2) |
| Living with partner, n (%) | 779 (7) |  | (25) Use of Ibuprofen, n (%) | 192 (2) |
| **(4) Alcohol drinking** (drinks/d) | 1 (0, 5) |  | (26) Use of Opium, n (%) | 160 (2) |
| **(5) Alcohol drinking groups** |  |  | (27) Use of Statin, n (%) | 2246 (21) |
| Non-drinker, n (%) | 2239 (21) |  | (28) Use of Valsartan, n (%) | 266 (2) |
| Moderate drinker, n (%) | 3128 (29) |  | **Dietary intakes** |  |
| Heavy drinker, n (%) | 5294 (50) |  | (29) Special diet, % | 1767 (17) |
| **(6) Occupation** |  |  | (30) Use of dietary supplements, % | 5100 (48) |
| Non-worker, n (%) | 4217 (40) |  | (31) Processed meat (g/d) | 13.5 (0, 78.2) |
| Part-time worker, n (%) | 1277 (12) |  | (32) Poultry (g/d) | 27.5 (0, 120.6) |
| Full-time worker, n (%) | 5167 (48) |  | (33) Fruits (cups/d) | 0.4 (0, 1.9) |
| **(7) Vigorous or moderate activity**, n (%) | 5442 (51) |  | (34) Vegetables (cups/d) | 1.3 (0.4, 2.9) |
| **(8) Sedentary lifestyle** (minutes/d) ^†^ | 360 (120, 600) |  | (35) Seafood (g/d) | 0 (0, 72.9) |
| **(9) Sleep** |  |  | (36) Whole grain (g/d) | 13.7 (0, 64.6) |
| ≤4 hours/night, n (%) | 588 (6) |  | (37) Eggs (g/d) | 6.8 (0, 43.9) |
| 5-8 hours/night, n (%) | 707 (7) |  | (38) Nuts and seeds (g/d) | 0.1 (0, 60.1) |
| ≥9 hours/night, n (%) | 9366 (88) |  | (39) Legumes (g/d) | 0 (0, 49.9) |
| **(10) Annual family income** |  |  | (40) Total diary (cups/d) | 1.2 (0.2, 3.0) |
| $ 0 to $14,999, n (%) | 1571 (15) |  | (41) Carbohydrates (g/d) | 232.5 (134.0, 380.2) |
| $15,000 to $34,999, n (%) | 2761 (26) |  | (42) Dietary fiber (g/d) | 15.2 (7.4, 28.1) |
| $35,000 to $64,999, n (%) | 2651 (25) |  | (43) SFAs (g/d) | 22.7 (10.8, 41.6) |
| $65,000 and over, n (%) | 3678 (34) |  | (44) MUFAs (g/d) | 25.6 (12.7, 46.1) |
| **(11) Socioeconomic status (PIR)** ^†^ | 2.51 (0.73, 5) |  | (45) PUFAs (g/d) | 16.2 (7.6, 30.3) |
| **(12) BMI** (kg/m^2^) | 28.4 (21.9, 38.5) |  | (46) Cholesterol (mg/d) | 246.0 (103.0, 535.5) |
| **(13) BMI groups** |  |  | (47) Magnesium (mg/d) | 275.5 (160.0, 457.0) |
| Healthy Weight, n (%) | 2780 (26) |  |  |  |
| Obesity, n (%) | 4249 (40) |  |  |  |
| Overweight, n (%) | 3509 (33) |  |  |  |
| Underweight, n (%) | 123 (1) |  |  |  |

*: Data presented as numbers and proportions or as medians (10th percentile, 90th percentile).

†: Systolic blood pressure, sedentary lifestyle, and socioeconomic status were categorized into fifth in the main analysis.

### Supplement Table 6: Results for testing the correlation between Schoenfeld residuals and ranked failure times to examine proportional hazard assumption violations for a specification that yielded a significant beneficial effect (HR:0.69, 95% CI:0.48-0.99, pvalue:0.04) of unprocessed red meat consumption on all-cause mortality. The specification used a standard model, continuous red meat, female sex, 60-79 years old, and variables in this table. A global test of proportional hazard assumption is also reported.

|  | CHISQ | DF | P value | Index |
| --- | --- | --- | --- | --- |
| Unprocessed red meat standard continuous | 1.59 | 1 | 0.21 |  |
| Marital status | 4.11 | 5 | 0.53 | 3 |
| Alcohol continuous | 0.47 | 1 | 0.49 | 4 |
| Sleep | 1.35 | 2 | 0.51 | 9 |
| Family income | 0.48 | 3 | 0.92 | 10 |
| Socioeconomic status | 7.56 | 4 | 0.11 | 11 |
| BMI Group | 0.85 | 3 | 0.84 | 13 |
| History of hypercholesterolemia | 0.02 | 1 | 0.89 | 16 |
| History of hypertension | 0.05 | 1 | 0.83 | 17 |
| History of depression | 0.34 | 1 | 0.56 | 19 |
| History of cardiovascular disease | 2.40 | 1 | 0.12 | 20 |
| History of cancer or malignancy | 0.88 | 1 | 0.35 | 21 |
| Family history of myocardial infarction | 1.89 | 1 | 0.17 | 23 |
| Use of Aspirin | 1.05 | 1 | 0.31 | 24 |
| Use of Ibuprofen | 1.36 | 1 | 0.24 | 25 |
| Use of Statin | 0.24 | 1 | 0.62 | 27 |
| Use of dietary supplement | 0.02 | 1 | 0.88 | 30 |
| Poultry | 0.07 | 1 | 0.79 | 32 |
| Fruits | 0.01 | 1 | 0.90 | 33 |
| Whole grain | 0.68 | 1 | 0.41 | 36 |
| Eggs | 0.36 | 1 | 0.55 | 37 |
| Nuts seeds | 4.22 | 1 | 0.04 | 38 |
| Legumes | 0.85 | 1 | 0.36 | 39 |
| Total dairy | 4.26 | 1 | 0.04 | 40 |
| Carbohydrates | 7.06 | 1 | 0.01 | 41 |
| Dietary fiber | 0.39 | 1 | 0.53 | 42 |
| Monounsaturated fatty acid | 2.33 | 1 | 0.13 | 44 |
| Magnesium | 4.23 | 1 | 0.04 | 47 |
| Cohort year | 2.71 | 3 | 0.44 |  |
| Age continuous | 0.08 | 1 | 0.78 |  |
| Smoking | 0.13 | 2 | 0.94 |  |
| Total energy | 4.58 | 1 | 0.03 |  |
| Menopausal status | 0.00 | 1 | 0.97 |  |
| Hormone therapy use | 1.75 | 1 | 0.19 |  |
| Parity | 0.86 | 1 | 0.36 |  |
| Oral contraceptive use | 0.10 | 1 | 0.76 |  |
| Global | 58.41 | 51 | 0.22 |  |

### Supplement Table 7: Results for testing the correlation between Schoenfeld residuals and ranked failure times to examine proportional hazard assumption violations for a specification that yielded a non-significant beneficial effect (HR:0.74, 95% CI:0.45-1.22, pvalue:0.24) of unprocessed red meat consumption on all-cause mortality. The specification used a multivariable nutrient density model, quartile red meat, male sex, 40-59 years old, and variables in this table. A global test of proportional hazard assumption is also reported.

|  | CHISQ | DF | P value | Index |
| --- | --- | --- | --- | --- |
| Unprocessed red meat density quartile | 1.73 | 3 | 0.63 |  |
| Education | 5.14 | 4 | 0.27 | 2 |
| Marital status | 3.72 | 5 | 0.59 | 3 |
| Alcohol continuous | 2.51 | 1 | 0.11 | 4 |
| Activity | 1.93 | 1 | 0.16 | 7 |
| Sedentary lifestyle | 2.43 | 4 | 0.66 | 8 |
| Socioeconomic status | 6.00 | 4 | 0.20 | 11 |
| BMI Group | 1.24 | 3 | 0.74 | 13 |
| General health condition | 6.43 | 4 | 0.17 | 15 |
| History of hypertension | 0.22 | 1 | 0.64 | 17 |
| History of diabetes | 0.00 | 1 | 0.99 | 18 |
| History of depression | 0.06 | 1 | 0.80 | 19 |
| History of cardiovascular disease | 4.68 | 1 | 0.03 | 20 |
| History of cancer or malignancy | 0.26 | 1 | 0.61 | 21 |
| Use of Opium | 0.02 | 1 | 0.88 | 26 |
| On special diet | 0.48 | 1 | 0.49 | 29 |
| Dietary supplement | 0.02 | 1 | 0.89 | 30 |
| Fruits | 0.08 | 1 | 0.77 | 33 |
| Vegetables | 6.66 | 1 | 0.01 | 34 |
| Seafood | 0.61 | 1 | 0.44 | 35 |
| Saturated fat | 0.32 | 1 | 0.57 | 43 |
| Polyunsaturated fatty acid | 0.25 | 1 | 0.62 | 45 |
| Cohort year | 2.54 | 3 | 0.47 |  |
| Age continuous | 1.84 | 1 | 0.17 |  |
| Smoking | 0.33 | 2 | 0.85 |  |
| Total energy | 0.26 | 1 | 0.61 |  |
| Global | 50.78 | 49 | 0.40 |  |

### Supplement Table 8: Results for testing the correlation between Schoenfeld residuals and ranked failure times to examine proportional hazard assumption violations for a specification that yielded a significant beneficial effect (HR:0.56, 95% CI:0.31-0.99, pvalue:0.05) of unprocessed red meat consumption on all-cause mortality. The specification used a multivariable nutrient density model, quintile red meat, male sex, 40-59 years old, and variables in this table. A global test of proportional hazard assumption is also reported.

|  | CHISQ | DF | P value | Index |
| --- | --- | --- | --- | --- |
| Unprocessed red meat density quintiles | 6.19 | 4 | 0.19 |  |
| Race Ethnicity | 9.06 | 4 | 0.06 | 1 |
| Alcohol continuous | 2.31 | 1 | 0.13 | 4 |
| Sleep | 4.05 | 2 | 0.13 | 9 |
| Socioeconomic status | 5.86 | 4 | 0.21 | 11 |
| BMI continuous | 1.32 | 1 | 0.25 | 12 |
| Systolic blood pressure | 0.26 | 4 | 0.99 | 14 |
| General health condition | 5.35 | 4 | 0.25 | 15 |
| History of hypertension | 0.07 | 1 | 0.80 | 17 |
| Family history of myocardial infarction | 0.40 | 1 | 0.53 | 23 |
| Use of Aspirin | 2.34 | 1 | 0.13 | 24 |
| Use of Ibuprofen | 0.82 | 1 | 0.37 | 25 |
| Use of Opium | 0.14 | 1 | 0.70 | 26 |
| Use of Valsartan | 0.76 | 1 | 0.38 | 28 |
| On special diet | 1.00 | 1 | 0.32 | 29 |
| Processed meat | 0.28 | 1 | 0.59 | 31 |
| Seafood | 0.60 | 1 | 0.44 | 35 |
| Whole grain | 0.03 | 1 | 0.87 | 36 |
| Eggs | 0.28 | 1 | 0.59 | 37 |
| Nuts seeds | 0.03 | 1 | 0.86 | 38 |
| Legumes | 0.03 | 1 | 0.85 | 39 |
| Total dairy | 0.00 | 1 | 0.99 | 40 |
| Carbohydrates | 2.18 | 1 | 0.14 | 41 |
| Monounsaturated fatty acid | 0.40 | 1 | 0.53 | 44 |
| Polyunsaturated fatty acid | 0.27 | 1 | 0.61 | 45 |
| Cohort year | 2.58 | 3 | 0.46 |  |
| Age continuous | 1.91 | 1 | 0.17 |  |
| Smoking | 0.25 | 2 | 0.88 |  |
| Total energy | 0.27 | 1 | 0.60 |  |
| Global | 62.53 | 48 | 0.08 |  |

### Supplement Table 9: Results for testing the correlation between Schoenfeld residuals and ranked failure times to examine proportional hazard assumption violations for a specification that yielded a non-significant harmful effect (HR:1.03, 95% CI:0.62-1.72, pvalue:0.90) of unprocessed red meat consumption on all-cause mortality. The specification used a standard model, continuous red meat, female sex, 40-59 years old, and variables in this table. A global test of proportional hazard assumption is also reported.

|  | CHISQ | DF | P value | Index |
| --- | --- | --- | --- | --- |
| Unprocessed red meat standard continuous | 1.94 | 1 | 0.16 |  |
| Education | 4.37 | 4 | 0.36 | 2 |
| Marital status | 3.88 | 5 | 0.57 | 3 |
| Alcohol continuous | 0.75 | 1 | 0.39 | 4 |
| Activity | 2.81 | 1 | 0.09 | 7 |
| Sedentary lifestyle | 1.68 | 4 | 0.79 | 8 |
| Socioeconomic status | 7.95 | 4 | 0.09 | 11 |
| BMI Group | 3.44 | 3 | 0.33 | 13 |
| General health condition | 7.78 | 4 | 0.10 | 15 |
| History of hypertension | 0.87 | 1 | 0.35 | 17 |
| History of diabetes | 0.72 | 1 | 0.40 | 18 |
| History of depression | 0.00 | 1 | 0.98 | 19 |
| History of cardiovascular disease | 1.31 | 1 | 0.25 | 20 |
| History of cancer or malignancy | 3.48 | 1 | 0.06 | 21 |
| Use of Opium | 2.29 | 1 | 0.13 | 26 |
| On special diet | 1.24 | 1 | 0.27 | 29 |
| Dietary supplement | 0.96 | 1 | 0.33 | 30 |
| Fruits | 0.08 | 1 | 0.78 | 33 |
| Vegetables | 0.20 | 1 | 0.65 | 34 |
| Seafood | 0.48 | 1 | 0.49 | 35 |
| Saturated fat | 2.22 | 1 | 0.14 | 43 |
| Polyunsaturated fatty acid | 0.01 | 1 | 0.93 | 45 |
| Cohort year | 3.34 | 3 | 0.34 |  |
| Age continuous | 0.57 | 1 | 0.45 |  |
| Smoking | 0.84 | 2 | 0.66 |  |
| Total energy | 1.67 | 1 | 0.20 |  |
| Menopausal status | 1.21 | 1 | 0.27 |  |
| Hormone therapy use | 0.00 | 1 | 0.96 |  |
| Parity | 0.99 | 1 | 0.32 |  |
| Oral contraceptive use | 0.01 | 1 | 0.94 |  |
| Global | 51.99 | 51 | 0.44 |  |

### Supplement Table 10: Results for testing the correlation between Schoenfeld residuals and ranked failure times to examine proportional hazard assumption violations for a specification that yielded a non-significant harmful effect (HR:1.10, 95% CI:0.30-3.96, pvalue:0.89) of unprocessed red meat consumption on all-cause mortality. The specification used a standard model, quintile red meat, all sex, 20-39 years old, and variables in this table. A global test of proportional hazard assumption is also reported.

|  | CHISQ | DF | P value | Index |
| --- | --- | --- | --- | --- |
| Unprocessed red meat standard quintiles | 1.41 | 4 | 0.84 |  |
| Race Ethnicity | 5.04 | 4 | 0.28 | 1 |
| Education | 5.08 | 4 | 0.28 | 2 |
| Alcohol Group | 1.67 | 2 | 0.43 | 5 |
| Activity | 0.01 | 1 | 0.91 | 7 |
| Sleep | 4.00 | 2 | 0.14 | 9 |
| Socioeconomic status | 2.97 | 4 | 0.56 | 11 |
| BMI continuous | 1.21 | 1 | 0.27 | 12 |
| Systolic blood pressure | 14.78 | 4 | 0.01 | 14 |
| Family history of diabetes | 0.00 | 1 | 0.97 | 22 |
| Family history of myocardial infarction | 0.11 | 1 | 0.73 | 23 |
| Use of Ibuprofen | 1.05 | 1 | 0.31 | 25 |
| On special diet | 0.03 | 1 | 0.86 | 29 |
| Dietary supplement | 0.25 | 1 | 0.62 | 30 |
| Processed meat | 0.45 | 1 | 0.50 | 31 |
| Poultry | 0.07 | 1 | 0.79 | 32 |
| Fruits | 0.22 | 1 | 0.64 | 33 |
| Whole grain | 0.02 | 1 | 0.90 | 36 |
| Eggs | 0.10 | 1 | 0.76 | 37 |
| Total dairy | 1.03 | 1 | 0.31 | 40 |
| Carbohydrates | 0.05 | 1 | 0.83 | 41 |
| Monounsaturated fatty acid | 1.27 | 1 | 0.26 | 44 |
| Cholesterol | 0.12 | 1 | 0.73 | 46 |
| Magnesium | 0.00 | 1 | 0.97 | 47 |
| Cohort year | 1.32 | 3 | 0.73 |  |
| Age continuous | 0.89 | 1 | 0.35 |  |
| Gender | 0.09 | 1 | 0.76 |  |
| Smoking | 0.35 | 2 | 0.84 |  |
| Total energy | 0.02 | 1 | 0.88 |  |
| Global | 46.92 | 49 | 0.56 |  |

### Supplement Figure 1: Results of primary studies identified from the systematic review reporting on the effect of red meat on all-cause mortality

The results of each analysis are presented as squares with 95% confidence intervals represented as horizontal lines. Effect estimates to the left of the vertical line of no effect suggest that red meat reduces the risk of all-cause mortality. The effect estimates are ordered based on their magnitude

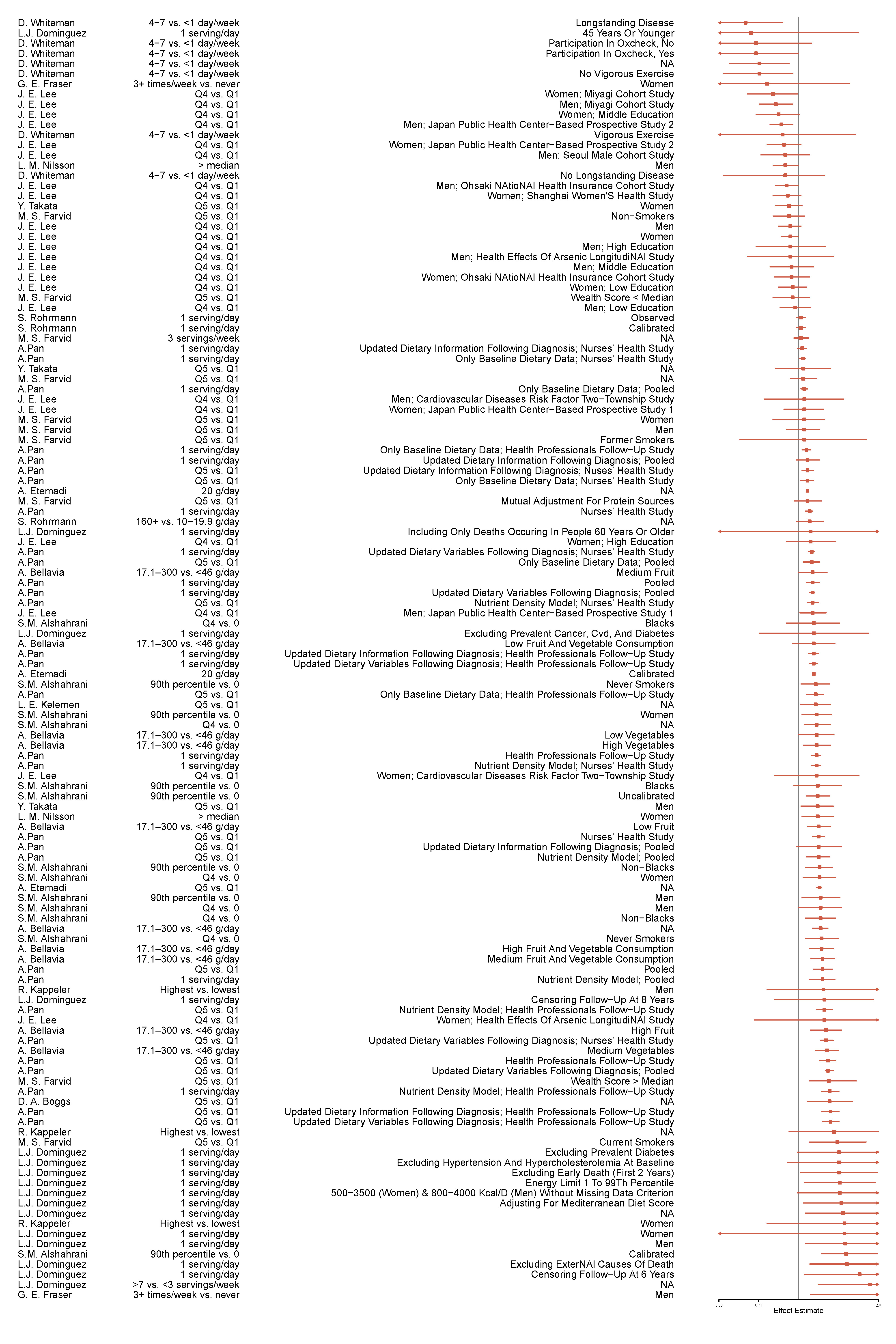

### Supplement Figure 2: Results of specification curve analysis stratified based on the operationalization of red meat in analytic models (e.g., continuous as 100 g/day, quartiles, and quintiles)

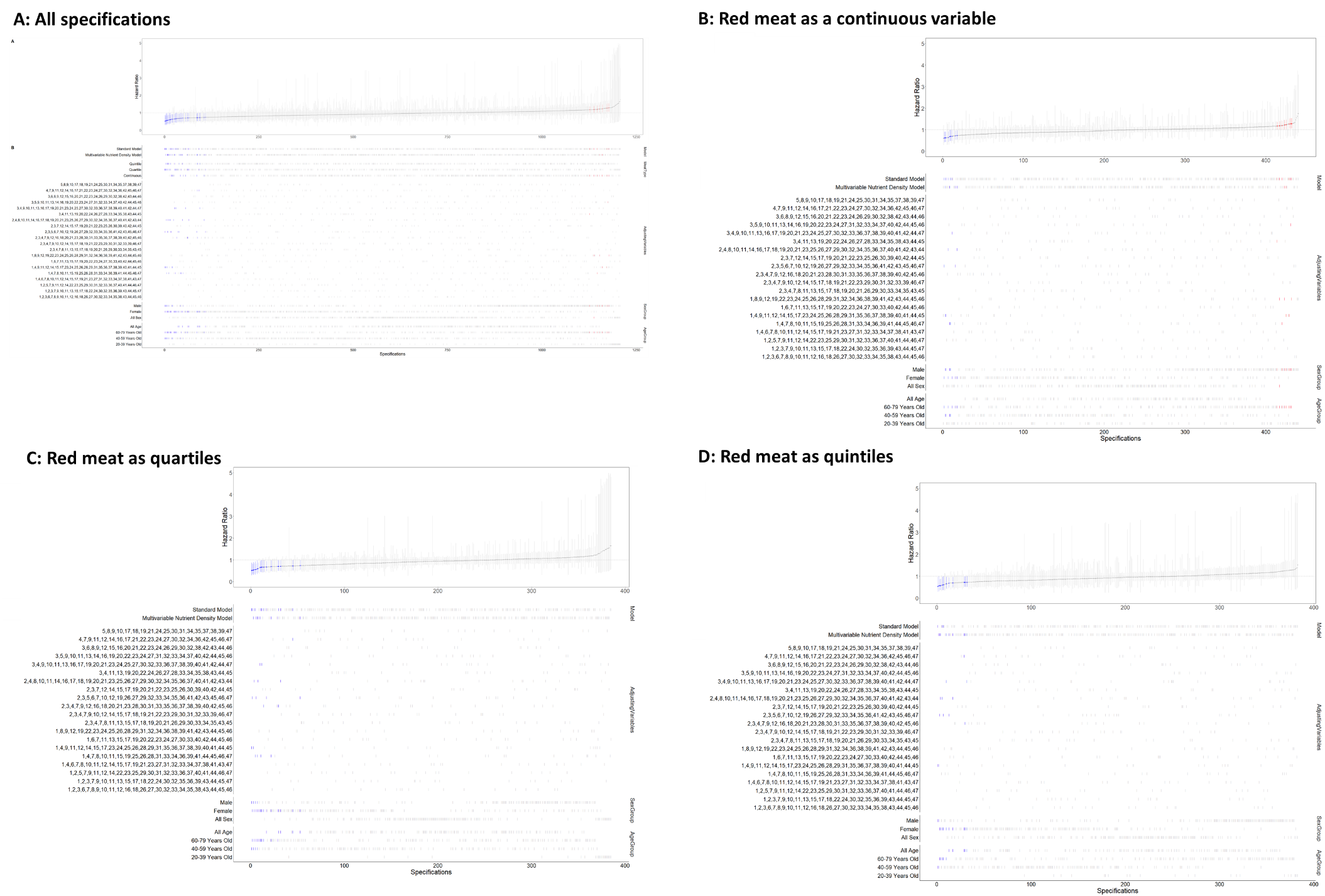

The upper portion of the four plots show HRs representing the effect of red meat on all-cause mortality. On the x axis are the unique analytic specifications. The y axis represents the magnitude of effect estimates. Each point on the graph represents the results of a unique analytic specification. Point estimates are shown in dark grey and 95% confidence intervals as light grey bars. Each point represents the results for the effect of red meat on all-cause mortality for a unique model. Point estimates are shown in dark grey and 95% confidence intervals as light grey bars. Each point represents the results for the effect of red meat on all-cause mortality for a unique model. Points in blue are statistically significant and suggest red meat to prevent all-cause mortality and points in red are statistically significant and indicate red meat to increase risk of all-cause mortality.

The lower parts of the plots show the characteristics of each analysis, including type of analytic model, operationalizations of variables, choice of covariates, and subgroups of interest. Each vertical line denotes the specific choice applied for each aspect of the analysis. We assigned a unique number to each covariate (Supplement 4 shows the number corresponding to each variable). Combinations of numbers in the graph represent combinations of covariates included in the model.

Panel A shows the results of a sample of all specifications and Panels B, C, and D show the results of specifications in which red meat is treated as a continuous variable, as quartiles, or as quintiles, respectively. For specifications in which red meat was treated as a continuous variable, we calculated HRs and associated confidence intervals corresponding to a 100 g/day increase in intake of red meat. For specifications in which red meat was treated as quantiles (e.g., quartiles or quintiles), we calculated hazard ratios and associated confidence intervals corresponding to the highest versus lowest quantile of red meat exposure.

### Supplement Figure 3: Plots for Schoenfeld residuals of covariates against ranked failure times to examine proportional hazard assumption violations for a specification that yielded a significant beneficial effect (HR:0.69, 95% CI:0.48-0.99, pvalue:0.04) of unprocessed red meat consumption on all-cause mortality. The specification used a standard model, continuous red meat, female sex, 60-79 years old, and variables in this table. P value=0.22 for the global test.

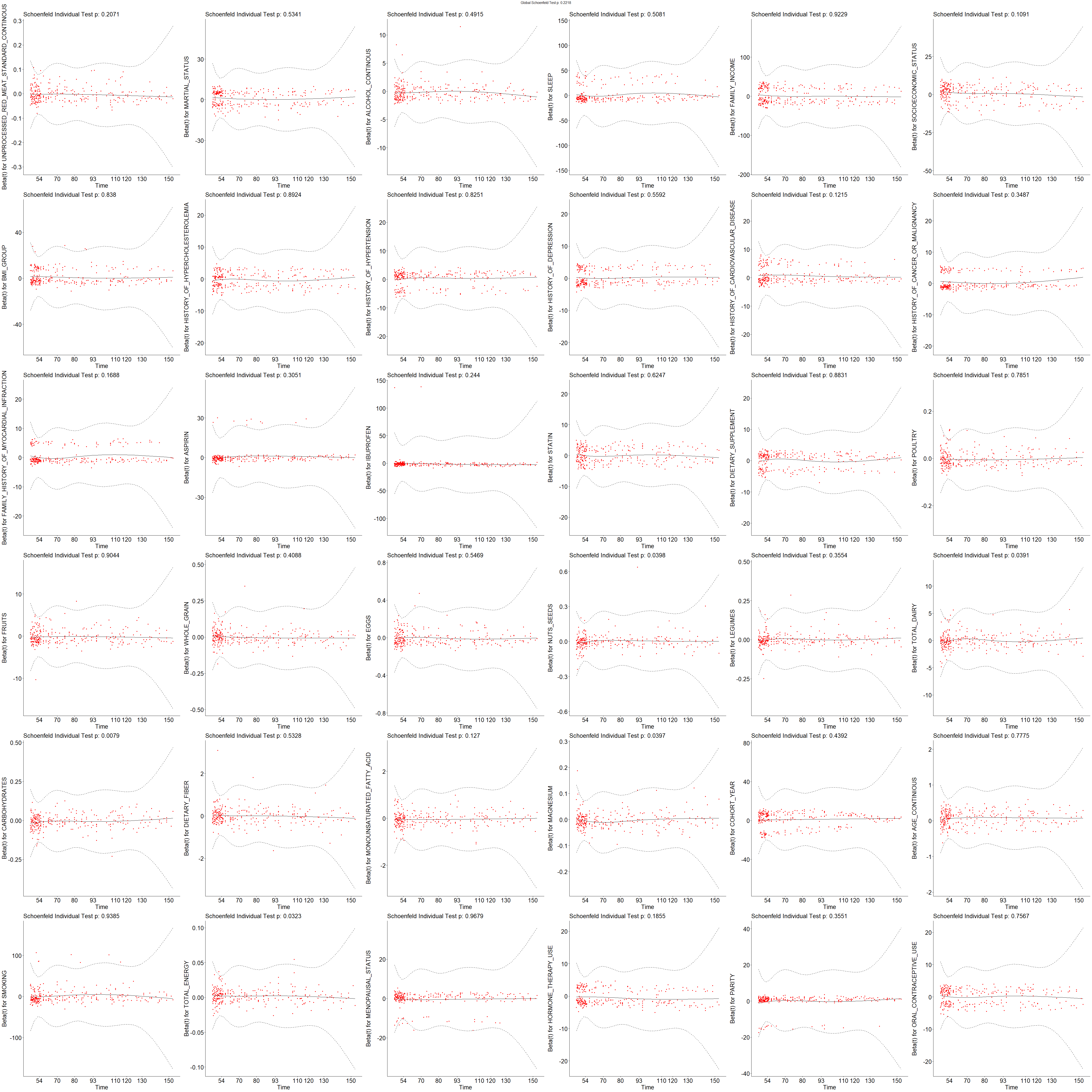

### Supplement Figure 4: Plots for Schoenfeld residuals of covariates against ranked failure times to examine proportional hazard assumption violations for a specification that yielded a non-significant beneficial effect (HR:0.74, 95% CI:0.45-1.22, pvalue:0.24) of unprocessed red meat consumption on all-cause mortality. The specification used a multivariable nutrient density model, quartile red meat, male sex, 40-59 years old, and variables in this table. P value=0.40 for the global test.

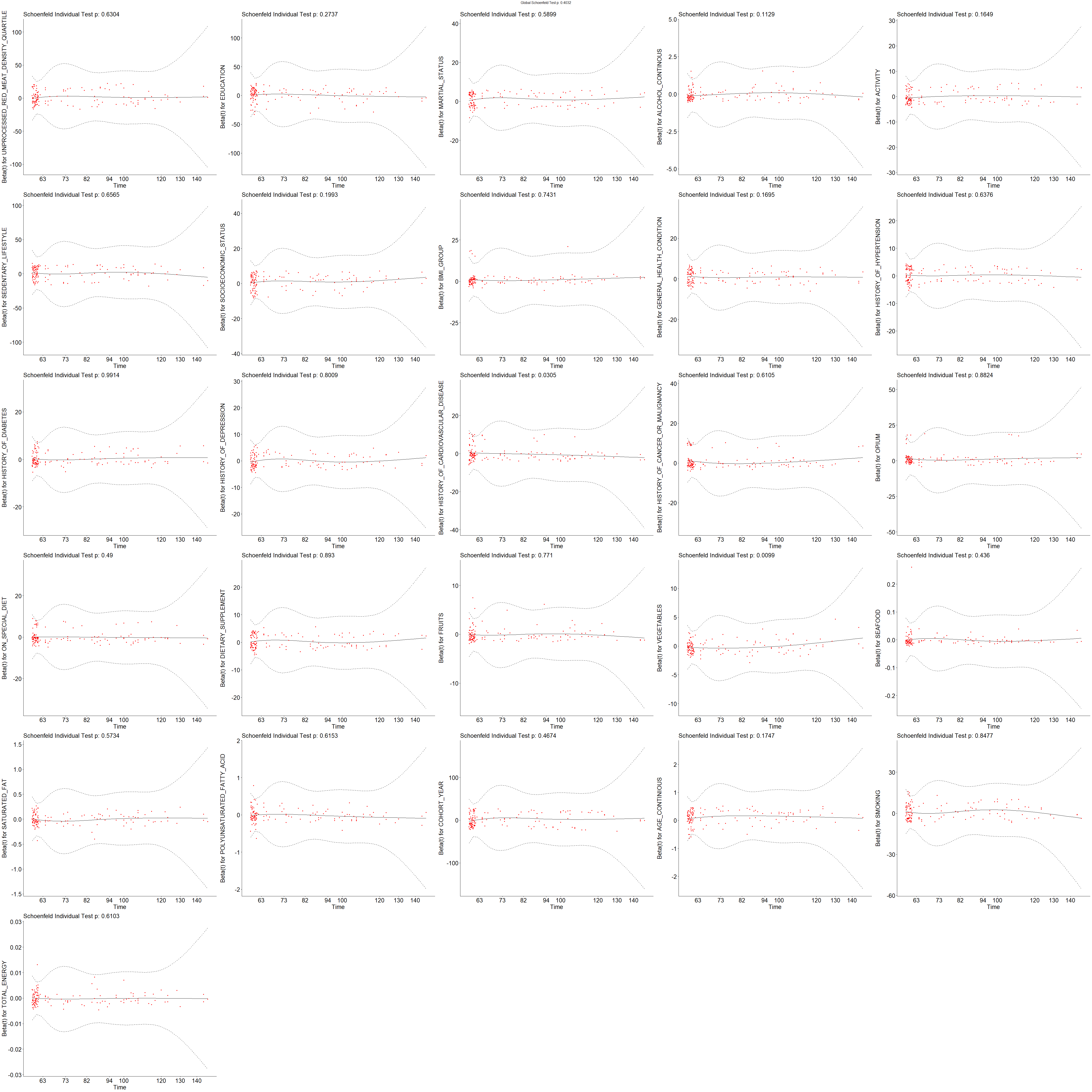

### Supplement Figure 5: Plots for Schoenfeld residuals of covariates against ranked failure times to examine proportional hazard assumption violations for a specification that yielded a significant beneficial effect (HR:0.56, 95% CI:0.31-0.99, pvalue:0.05) of unprocessed red meat consumption on all-cause mortality. The specification used a multivariable nutrient density model, quintile red meat, male sex, 40-59 years old, and variables in this table. P value =0.08 for the global test.

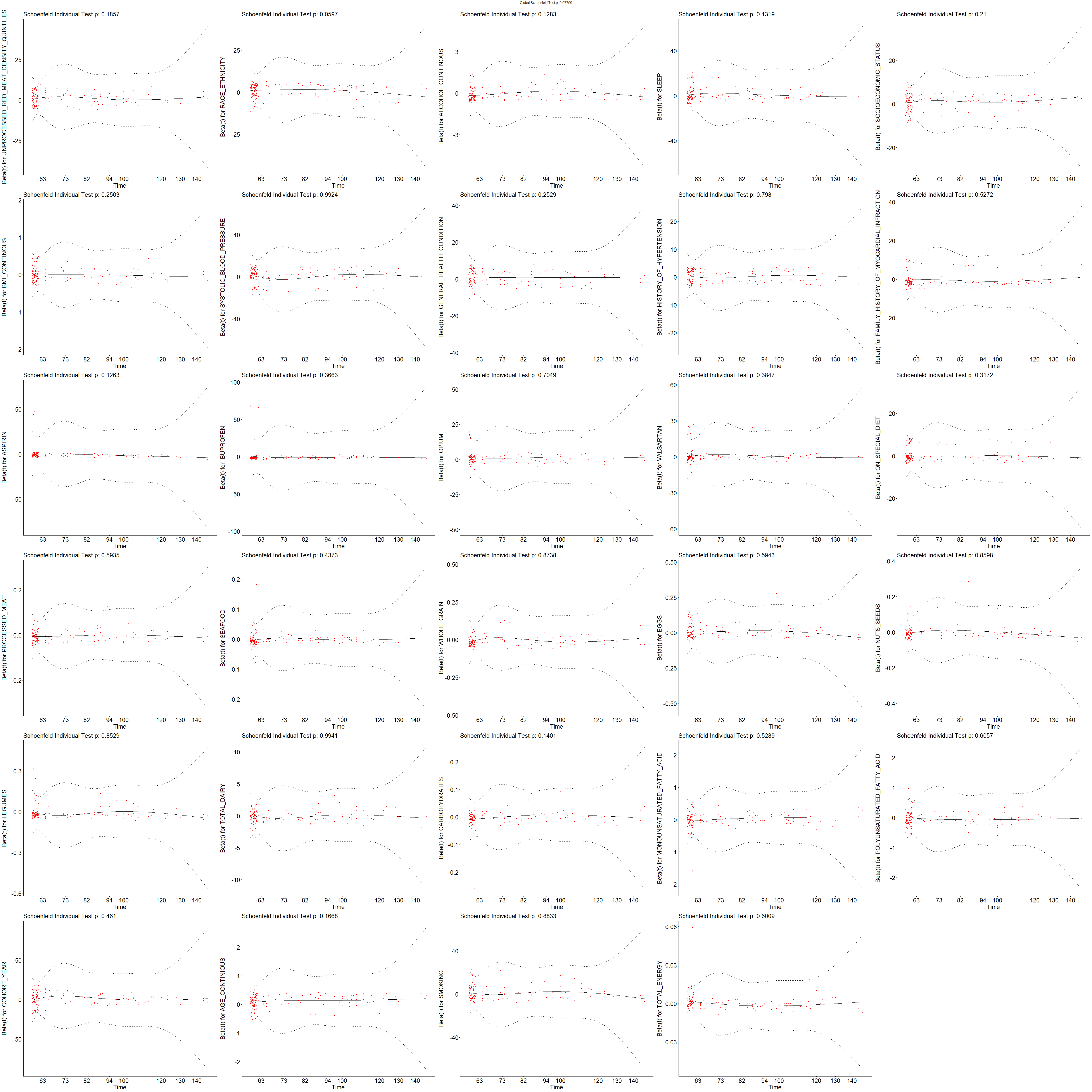

### Supplement Figure 6: Plots for Schoenfeld residuals of covariates against ranked failure times to examine proportional hazard assumption violations for a specification that yielded a non-significant harmful effect (HR:1.03, 95% CI:0.62-1.72, pvalue:0.90) of unprocessed red meat consumption on all-cause mortality. The specification used a standard model, continuous red meat, female sex, 40-59 years old, and variables in this table. P value=0.44 for the global test.

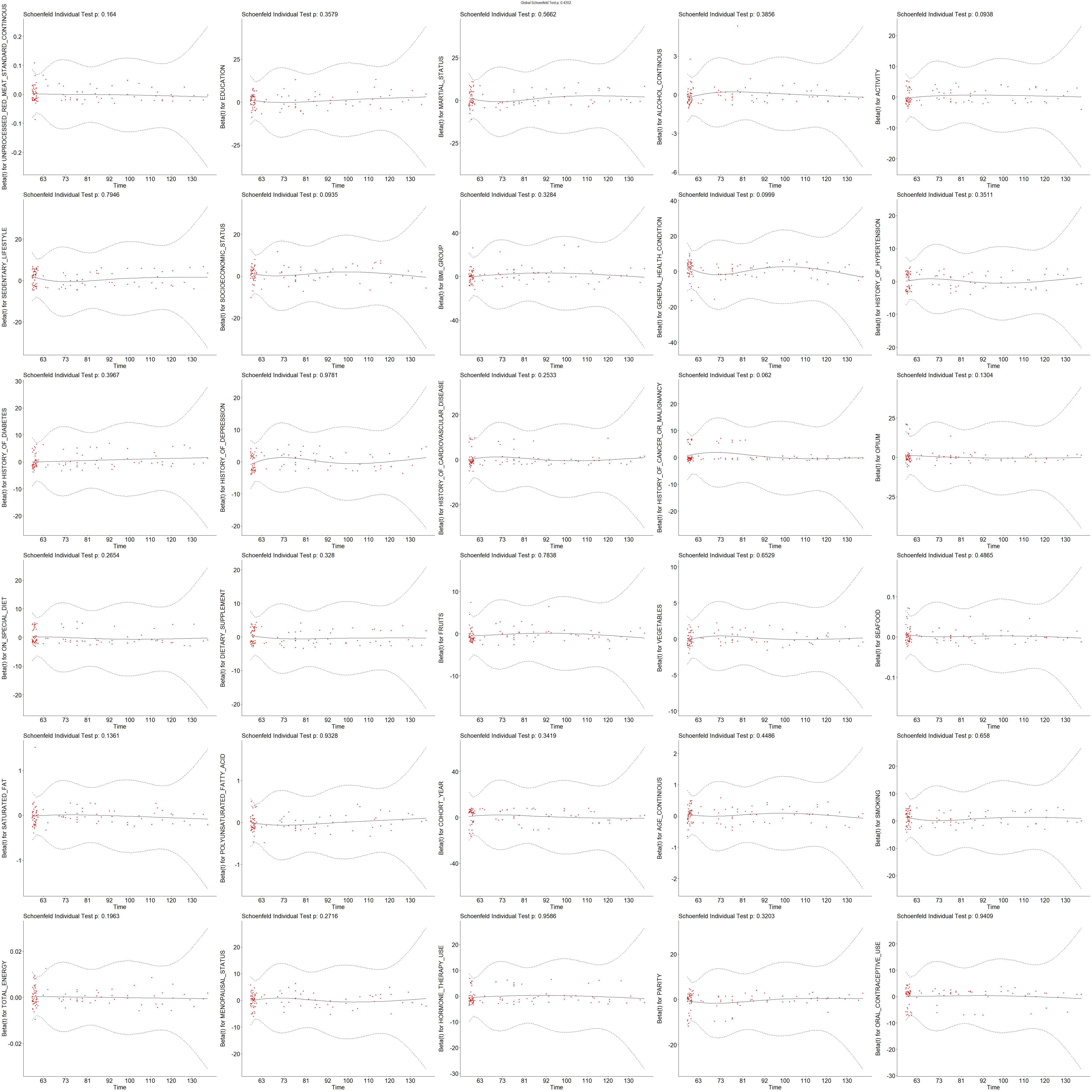

Supplement Figure 7: Plots for Schoenfeld residuals of covariates against ranked failure times to examine proportional hazard assumption violations for a specification that yielded a non-significant harmful effect (HR:1.10, 95% CI:0.30-3.96, pvalue:0.89) of unprocessed red meat consumption on all-cause mortality. The specification used a standard model, quintile red meat, all sex, 20-39 years old, and variables in this table. P value=0.56 for the global test.
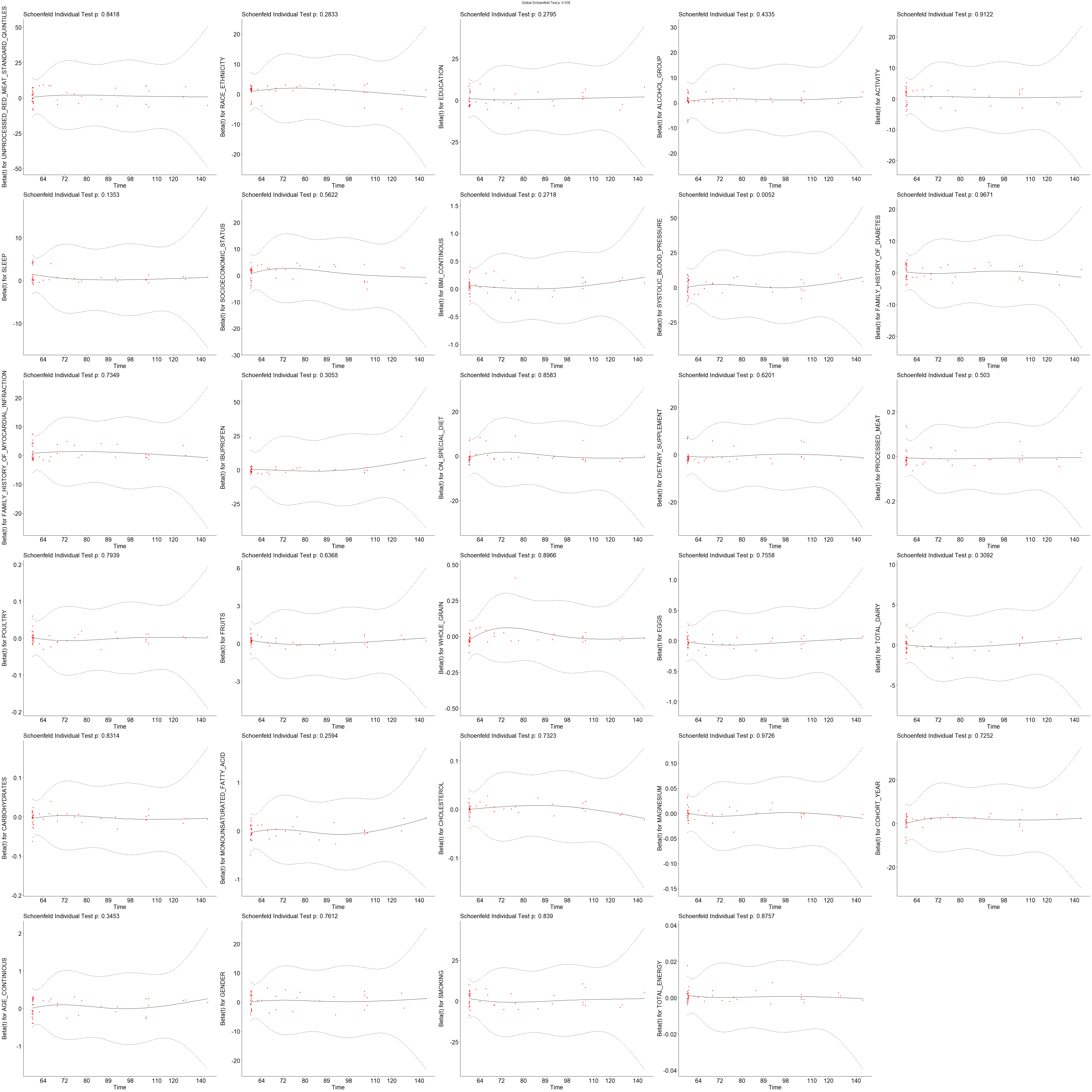
