## Supplementary figures and images for "Variations in the results of nutritional epidemiology studies due to analytic flexibility: Application of specification curve analysis to red meat and all-cause mortality"

### Supplement figure 1

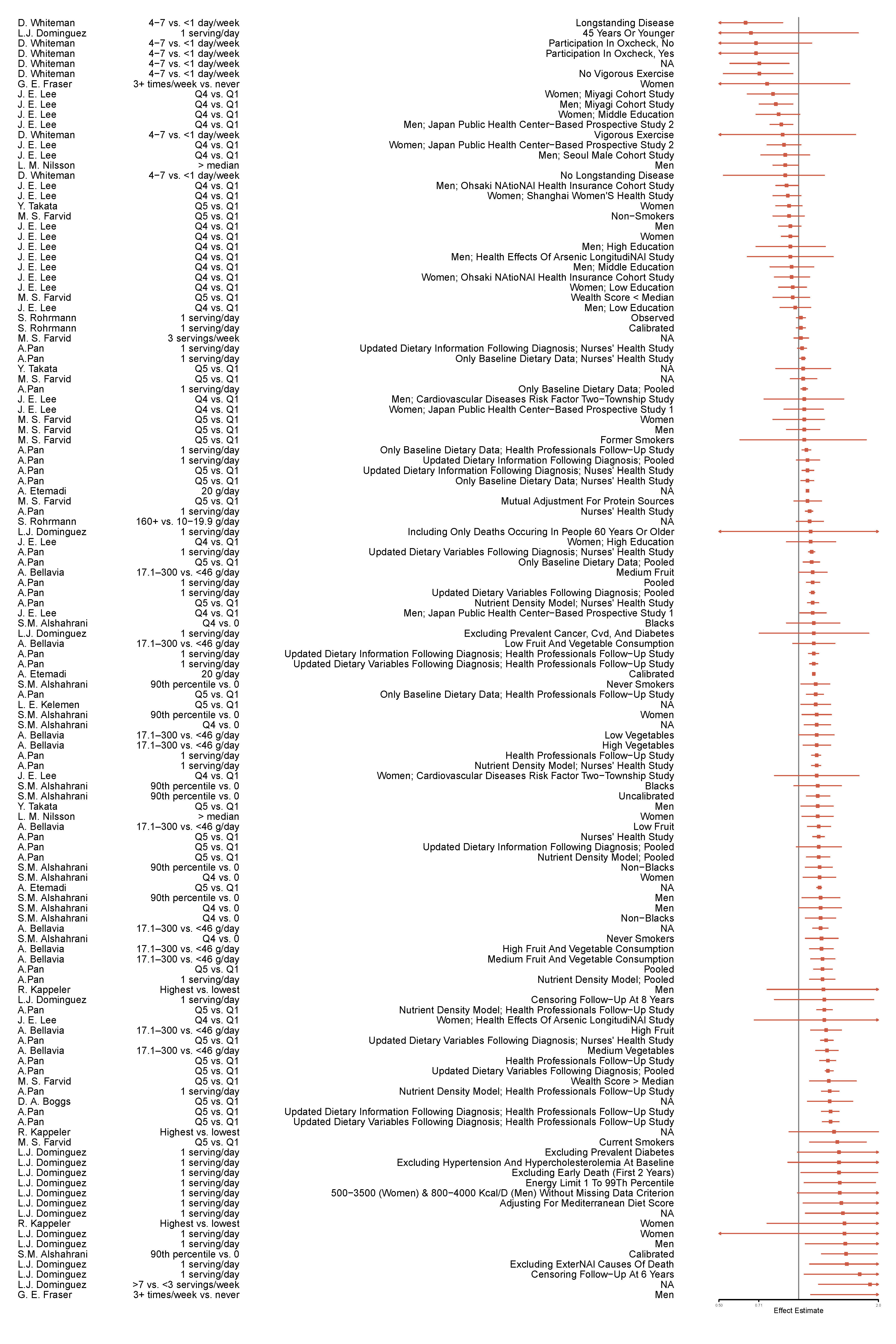

### Supplemental Figure 2B

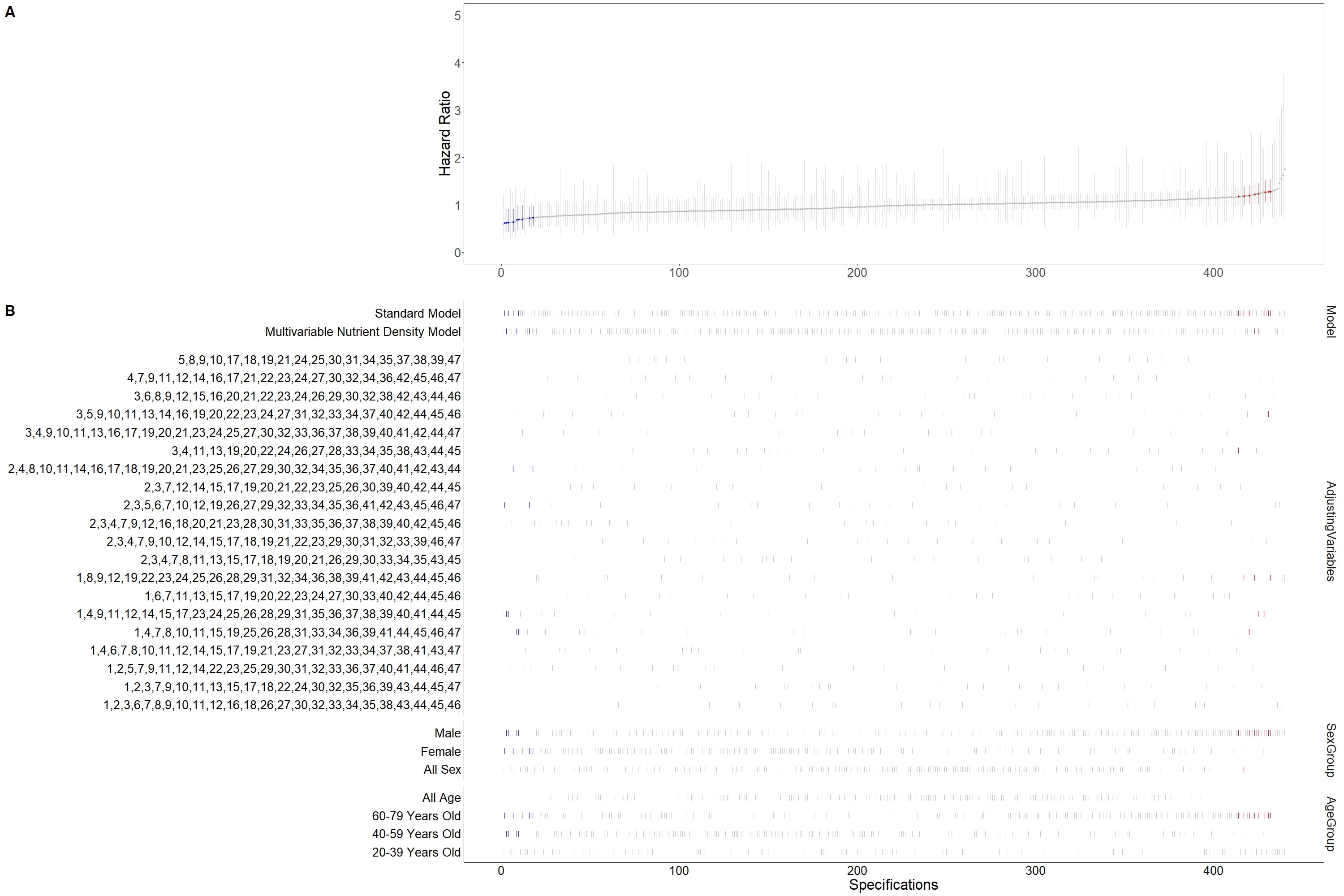

### Supplemental Figure 2C

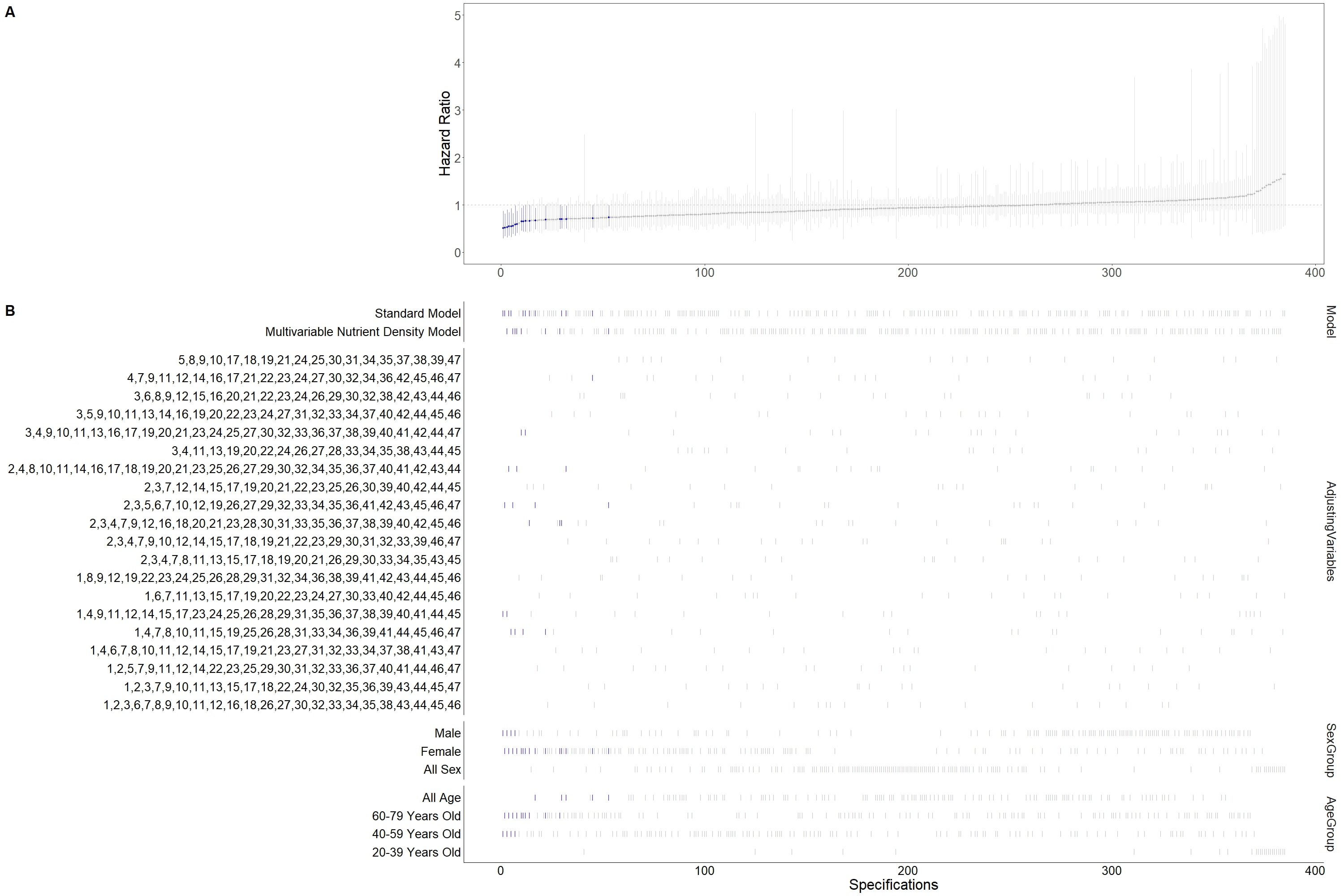

### Supplemental Figure 2D

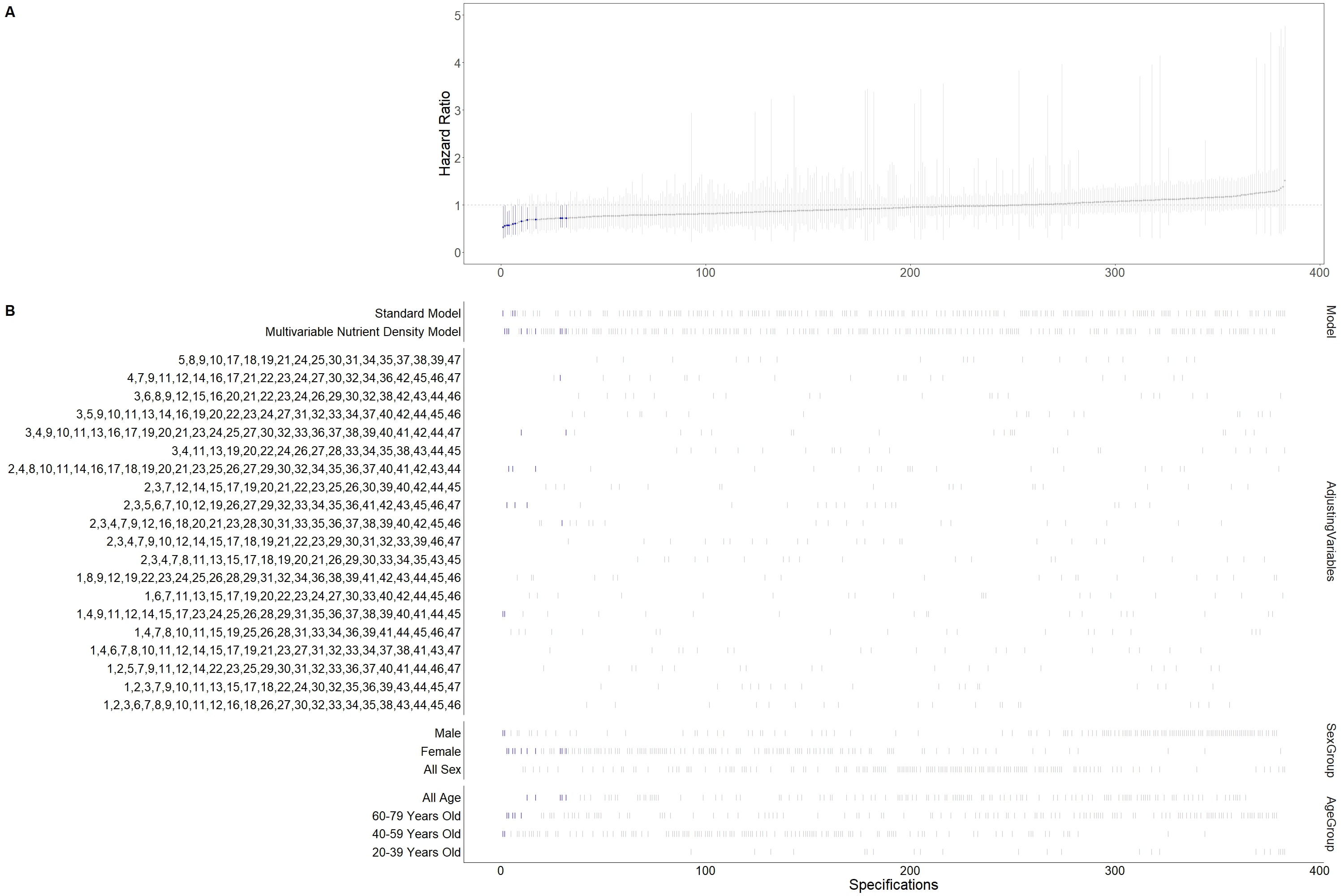
